# Exploring the genetic overlap between attention-deficit/hyperactivity disorder (ADHD) and migraine

**DOI:** 10.64898/2026.01.12.26343903

**Authors:** Pau Carabí-Gassol, Natalia Llonga, Silvia Alemany, Uxue Zubizarreta-Arruti, Manuel Donato Rodríguez-Romero, Valeria Macias-Chimborazo, Christian Fadeuilhe, Montse Corrales, Vanesa Richarte, Víctor J. Gallardo, Patricia Pozo-Rosich, Josep Antoni Ramos-Quiroga, Marta Ribasés, Judit Cabana-Domínguez, María Soler Artigas

## Abstract

**Background:** Attention-deficit/hyperactivity disorder (ADHD) and migraine are prevalent neurodevelopmental and neurological conditions, respectively, that contribute to individual disability and social burden. The biological mechanisms linking these disorders remain poorly understood.

**Methods:** We aimed to investigate their shared genetic architecture by integrating genomic data with a cross-trait analysis using the largest genome-wide association studies (GWAS) for ADHD and migraine to date. Variants were classified into concordant and discordant, according their direction of effect, and were followed-up with functional and colocalization analyses. Then, polygenic risk scores (PRSs) analyses were undertaken aiming to dissect their clinical heterogeneity, using in-house ADHD and migraine samples.

**Results:** We identified 961 pleiotropic SNPs across 29 loci, including 9 loci not previously related to either condition. Concordant variants were enriched in immune-related pathways, brain morphology, and autoimmune traits, while discordant variants mapped to genes associated with psychiatric, cardiovascular, and behavioural traits. Colocalization analysis showed more pleiotropic concordant loci than discordant loci (62% vs 17%). Individuals with higher PRS for migraine showed increased odds for ADHD. Also, migraine PRSs were positively associated to childhood headaches in individuals with ADHD. This association was stronger when restricting the PRS to concordant migraine-specific and pleiotropic variants, despite including only one quarter of them. Furthermore, ADHD individuals reporting childhood headaches also showed higher levels of anxiety, depression, neuroticism, and reduced cognitive performance in adulthood.

**Conclusions:** These findings suggest shared genetic mechanisms between ADHD and migraine, particularly involving neuroimmune and neurodevelopmental pathways, and support the utility of pleiotropy-informed PRSs models for understanding comorbid traits.

## 1. Introduction

Attention deficit/hyperactivity disorder (ADHD) is an early-onset neurodevelopment disorder affecting around 5% of children and 2.5% of adults (Faraone et al., 2015). It is characterized by impairing levels of hyperactivity, inattention, or impulsivity, which significantly impacts the academic, social, emotional and psychological functioning, resulting in high individual and society costs (Du Rietz et al., 2017). In adults, ADHD often presents a heterogenous clinical range of symptoms, extending beyond the typical paediatric motor symptoms, including broader functional impairments and emotional dysregulation (Katzman et al., 2017). Moreover, around 80% of the individuals diagnosed with ADHD have at least one coexisting psychiatric disorder (Sobanski et al., 2007). Non-psychiatric comorbidities are also frequent; systematic reviews suggest robust associations with obesity, asthma, sleep disorders, migraine and celiac diseases, with less consistent evidence for epilepsy, elimination disorders, and allergic or immunological disorders (Instanes et al., 2018).

A positive association between ADHD and migraine has been reported. Migraine is a common neurological disorder, with a prevalence around 11%, that usually starts in adolescence or early adulthood and is among the most disabling diseases worldwide (Gustavsson et al., 2011). The co-occurrence of ADHD and migraine has been well documented in adults (Hansen et al., 2018; Kittel-Schneider et al., 2022; Salem et al., 2018) and more recently confirmed in paediatric populations as well (Arruda et al., 2020). Additionally, the association between ADHD and migraine appears to be stronger in females, which may be partly explained by the higher prevalence of migraine in women compared to men (Arruda et al., 2020). ADHD co-occurrence with migraine has been related with two particular aspects of migraine, higher frequency and presence of visual symptoms (Arruda et al., 2020). While this comorbidity is well documented clinically and epidemiologically, the genetics and molecular mechanisms underlying the ADHD-migraine relationship remain poorly explored.

Both ADHD and migraine are complex, polygenic disorders influenced by genetic and environmental factors. Twin studies have estimated a heritability of around 72% for adults with ADHD (Larsson et al., 2014) and between 34% and 57% for migraine (Mulder et al., 2003). The largest genome-wide association study meta-analysis (GWAS-MA) of ADHD identified 27 loci and estimated that common variants account for about 14% of its heritability (Demontis et al., 2023). For migraine, the largest GWAS-MA to date identified 123 risk loci explaining approximately 10.6-11% of its heritability (Hautakangas et al., 2022). In 2018, the Brainstorm consortium published the first large-scale genomic analysis to identify shared genetic architecture between neurological and psychiatric disorders. In this study, migraine was the only neurological disease with a substantial genetic overlap with psychiatric disorders, including ADHD (Anttila et al., 2020). More recently, a study reported differences in polygenicity, with ADHD involving approximately four times the number of contributing variants compared to migraine (Ciochetti et al., 2025). The same study reported a genetic correlation of 20% between both disorders, and identified 19 pleiotropic loci (Anttila et al., 2020; Ciochetti et al., 2025). However, the approach used in this study did not account for differentiation between horizontal pleiotropy, where a genetic variant exerts direct effects on both traits, consistent with the presence of shared molecular mechanisms, and vertical pleiotropy, in which an effect on one trait influences the other through a causal relationship.

This study aimed to investigate the comorbidity between ADHD and migraine, by exploring shared molecular mechanisms and clinical heterogeneity, focusing on horizonal pleiotropy. To achieve this, we (i) performed a cross-trait analysis to identify shared genetic loci between ADHD and migraine, (ii) conducted downstream functional analyses to assess the biological relevance of the pleiotropic signals, and (iii) undertook polygenic risk score (PRS) analyses for ADHD, migraine, and pleiotropic variants to explore clinical heterogeneity.

## 2. Experimental procedures

### 2.1 Cross-trait analysis

#### 2.1.1 GWAS-MA samples and data processing

GWAS-MA summary statistics on ADHD included a total of 38,691 individuals with ADHD and 186,843 controls (Demontis et al., 2023) and for migraine, 48,975 individuals with migraine and 412,985 controls (Hautakangas et al., 2022); all of European ancestry. SNP quality control filtering is detailed in the *Supplementary Methods*.

#### 2.1.2 PolarMorphism

The PolarMorphism (von Berg et al., 2022) methodology was used to identify horizontal pleiotropic variants between ADHD and migraine using the summary statistics described above. PolarMorphism takes common SNPs (N=5,243,166), from the two studies and decorrelates the input data to attenuate vertical pleiotropy. Then, it takes decorrelated Z-scores from both traits as cartesian coordinates and transforms them into polar coordinates obtaining a measure of overall effect, (distance from the origin), and of sharedness between traits, theta (angle with the x-axis*, Supplementary Figure 1 A*). This method first selects SNPs with a significant overall effect (*r* false discovery rate (FDR) corrected p-value, q-value < 0.05), and then it tests whether these SNPs are also pleiotropic (*θ* q-value < 0.05).

Top hits from PolarMorphism were clumped to detect independent loci and compared with the results in the original GWAS-MA for ADHD or migraine, or in the recent cross-trait analysis between them to identified novel loci (Demontis et al., 2023; Hautakangas et al., 2022) (Ciochetti et. al. 2025).

Functional annotation and colocalization analyses were undertaken to follow-up cross-trait results, separately for variants with the same direction of effect in ADHAD and migraine (concordant) and with different direction of effect (discordant).

Details on SNP classification according to their direction of effect and value, as well as clumping, new loci identification and follow-up analyses can be found in the *Supplementary Methods*.

### 2.2 Polygenic risk scores analysis

#### 2.2.1 Discovery sample

The summary statistics described above were used as discovery datasets. For ADHD, because of sample overlap with our target sample, the effect of the Spanish subsample was removed from the original GWAS-MA summary statistics (Demontis et al., 2023), leaving 37,937 cases and 185,103 controls.

#### 2.2.2 Target sample

##### ADHD sample

An in-house clinical sample of 1,775 ADHD cases (68% males, mean age 28±13.3 years) was evaluated and recruited from a restricted geographic area in a specialized out-patient program for adult ADHD and by a single clinical group at Hospital Universitari Vall d’Hebron (HUVH) in Barcelona, Spain.

Clinical assessment of ADHD diagnosis, severity, psychiatric comorbidities and personality features was performed using structured interviews and self-reported questionnaires as previously described (Mortimer et al., 2020). Clinical assessment and exclusion criteria are detailed in the *Supplementary Methods*. All clinical data were curated, summarized and filtered by effective sample size >200, resulting in 35 clinical items included in the study (*Supplementary Table 1*), as well as two questions related to migraine or headache: (i) Adulthood migraine: “Since you were 18 years old, have you had migraine?”; (ii) Childhood headache: “Medical problems during childhood: headache”.

##### Migraine sample

An in-house clinical sample of 1,325 migraine cases (22.8% males, mean age 40±12.3) was retrospectively evaluated and recruited in the specialized Headache Clinic of HUVH in Barcelona, Spain. Participants were diagnosed with migraine according to the International Classification of Headache Disorders, 2nd edition (ICHD-II)(Olesen, 2004). For ADHD screening, participants were electronically contacted and invited to complete the Adult ADHD Self-Report Scale (ASRS v1.1)(Ramos Quiroga et al., 2009). Further information can be found in the *Supplementary Methods*.

##### Control sample

The control groups for the ADHD (n=2,558; 50% males, mean age 52±13.7) and migraine (n=1,664; 58.11% males, mean age 47±19.3) clinical samples consisted of blood donors. Individuals with ADHD symptomatology were excluded (details can be found in the *Supplementary Methods*).

The study was approved by the Clinical Research Ethics Committee (CREC) of Hospital Universitari Vall d’Hebron, methods were performed in accordance with the relevant guidelines and regulations and written informed consent was obtained from all subjects before inclusion in the study.

Details on genotyping and imputation are given in the *Supplementary Methods*.

#### 2.2.3 Association testing of polygenic risk scores

Three PRS were generated for ADHD, including all variants (PRS_ADHD_) and including ADHD-specific and migraine-ADHD pleiotropic variants, separately for concordant (PRS_ADHD_CC_) and discordant variants (PRS_ADHD_DC_). The same criteria were used to generate three PRS for migraine (PRS_Mig_, PRS_Mig_CC_ and PRS_Mig_DC_). Detailes can be found in the *Supplementary Methods*.

We tested the association between PRS_ADHD_ and ADHD, and between PRS_Mig_ and migraine, as positive controls. To assess cross-trait effects, we also tested whether PRS_ADHD_ was associated with migraine, and whether PRS_Mig_ was associated with ADHD. Logistic regression was used for all models with age, sex, and the first 10 PC as covariates. Additionally, in the model used to test the association with ADHD, GWAS wave was also included as covariate. Quintiles plots were constructed to assess the association across PRSs distributions comparing the first quintile with each of the subsequent quintiles.

##### PRS_Mig_ association with clinical heterogeneity in the ADHD sample

Clinical information was available for 939 individuals with ADHD (*Supplementary Table 1*). This clinical dataset was considered to test the association of PRSs for migraine (PRS_Mig_, PRS_Mig_CC_ and PRS_Mig_DC_) with adulthood migraine and childhood headache using logistic regression and age, sex, GWAS wave and the first 10 PC as covariates.

For the significant associations identified, the ADHD sample was stratified into two groups: individuals who answered affirmatively to the adulthood migraine/childhood headache question and those who answered negatively. In order to identify clinical characteristics that differed between the two groups, we compared the clinical profiles (details can be found in the *Supplementary Methods*).

Finally, we tested the association between the clinical items showing significant differences between the two groups and PRSs for migraine previously associated with adulthood migraine/childhood headache. Logistic regression, including age, sex, GWAS wave and the first 10 PC as covariables were used.

##### PRS_ADHD_ association with clinical heterogeneity in the migraine sample

For the subset of 256 individuals with migraine and information on ADHD screening according to ASRS 1.1, associations between PRSs for ADHD (PRS_ADHD_, PRS_ADHD_CC_ and PRS_ADHD_DC_) and ADHD were tested using logistic regression, including age, sex and the first 10 PC as covariates.

## 3. Results

### Cross-trait analysis

The cross-trait analysis between ADHD and migraine identified 961 pleiotropic SNPs distributed across 29 independent loci. A total of 32 independent lead SNPs were detected after clumping, 17 of them showing concordant direction of effects while 15 showed discordant direction of effects (*Supplementary Table 2, Table 1, **Error! Reference source not found.**1*). Out of these 32 independent lead SNPs, 9 novel variants not previously identified in the original GWAS-MA of ADHD or migraine neither in the previous cross-trait analysis done by Ciochetti et. al. (Ciochetti et al., 2025; Demontis et al., 2023; Hautakangas et al., 2022). Among the new lead SNPs identified, two were concordant and seven were discordant variants *(Table 1).* Functional annotation on the subset of concordant variants, which included 17 lead SNPs and the SNPs in LD with them (r^2^>0.6; N=1,958; *Supplementary Table 3),* revealed that 89.90% of these variants were located in euchromatin regions, with most signals intronic (61.33%), intergenic (14.7%) or non-coding RNA intronic (14.44%) (*Supplementary Figure 2A*). A total of 70% of this set of SNPs (n=1373) were eQTL for at least one gene in brain tissue, according to GTEx v8 and BRAINEAC (*Supplementary Table 3)*. For the discordant variants, comprising 15 lead variants and those in LD with them (N=1504 variants, *Supplementary Table 4*), functional annotation revealed that 54.45% of these variants lay on euchromatin, and most of the signals were intronic (19.28%), intergenic (62.29%) or non-coding RNA intronic (9.44%) (*Supplementary Figure 2B*). A total of 22% of the discordant SNPs (n=303) were eQTL for at least one gene on brain area, according to GTEx v8 and BRAINEAC (*Supplementary Table 4*).

Concordant variants mapped to 83 genes, enriched for neurological and brain structure traits (i.e., brain morphology, subcortical volume, white matter integrity), cerebrovascular disorders (i.e., lacunar stroke, non-lobar intracerebral haemorrhage), as well as traits related to the immune system (i.e., antigen processing and presentation, natural killer cell mediated cytotoxicity) and autoimmune and inflammatory diseases (such as vitiligo, asthma or COPD) among others (*Supplementary Table 5A*). The enriched sets related to immune system were mainly driven by five genes (*RAET1E, RAET1G, RAET1L, ULBP1* and *ULBP2, Supplementary Table 5A*). On the other hand, the 48 genes mapped by discordant variants were enriched for blood and metabolic traits (microalbuminuria), cardiovascular traits (i.e., mean arterial pressure, systolic blood pressure, coronary artery diseases), neurological and sensory conditions (i.e., migraine, headaches), lifestyle and behavioral traits (i.e., age at first sexual intercourse, number of sexual partners, smoking initiation), and mental health disorders (i.e., autism or schizophrenia, bipolar disorder, ADHD; *Supplementary Tables 5B*). Some neurological and brain structure traits (i.e., brain morphology evaluated with MOSTest, anorexia nervosa and refractive error) were enriched for both the concordant and discordant set, however, there was no overlap of genes (*Supplementary Tables 5*).

After colocalization analysis on the 32 lead SNPs identified in the cross-trait analysis, 25 had posterior probability (PP)>0.6 for at least one of the five tested hypotheses. Of those SNPs, in the concordant set 8 out of 13 (62%) showed pleiotropy (PP>0.6 for the H_3_ or H_4_ hypothesis), five of which colocalized (PP>0.6 for H_4_), and three showed evidence of association only with one disorder (PP>0.6 for H_1_ or H_2_; *Supplementary Table 6A*). In contrast, within the discordant set, only 2 out of 12 (17%) showed evidence of pleiotropy, both of them colocalizing (PP>0.6 for H_4_), and 10 (83%) showed evidence of association only with one disorder (PP>0.6 for H_1_ or H_2_; *Supplementary Table 6B*). Colocalization plots are provided in the *Supplementary Figures* (*Supplementary Figures 3*).

### Polygenic risk score analysis

In the ADHD case-control clinical sample, PRS_ADHD_ was significantly associated with ADHD diagnosis (OR=1.20;95%CI=(1.10,1.32);P=5.61e-05), whereas PRS_Mig_ showed no significant association with ADHD (OR=1.05;95%CI=(0.98, 1.14);P=0.16). However, individuals in the highest PRS_Mig_ quintile showed a significant increase in odds for ADHD compared to the first quintile (OR=1.49;95%CI=(1.16,1.92);P=0.01), suggesting that higher genetic load for migraine may be associated with increased risk of developing ADHD (*Figure 2A; Supplementary table 7, Supplementary Figure 4)*.

**Figure 1.**
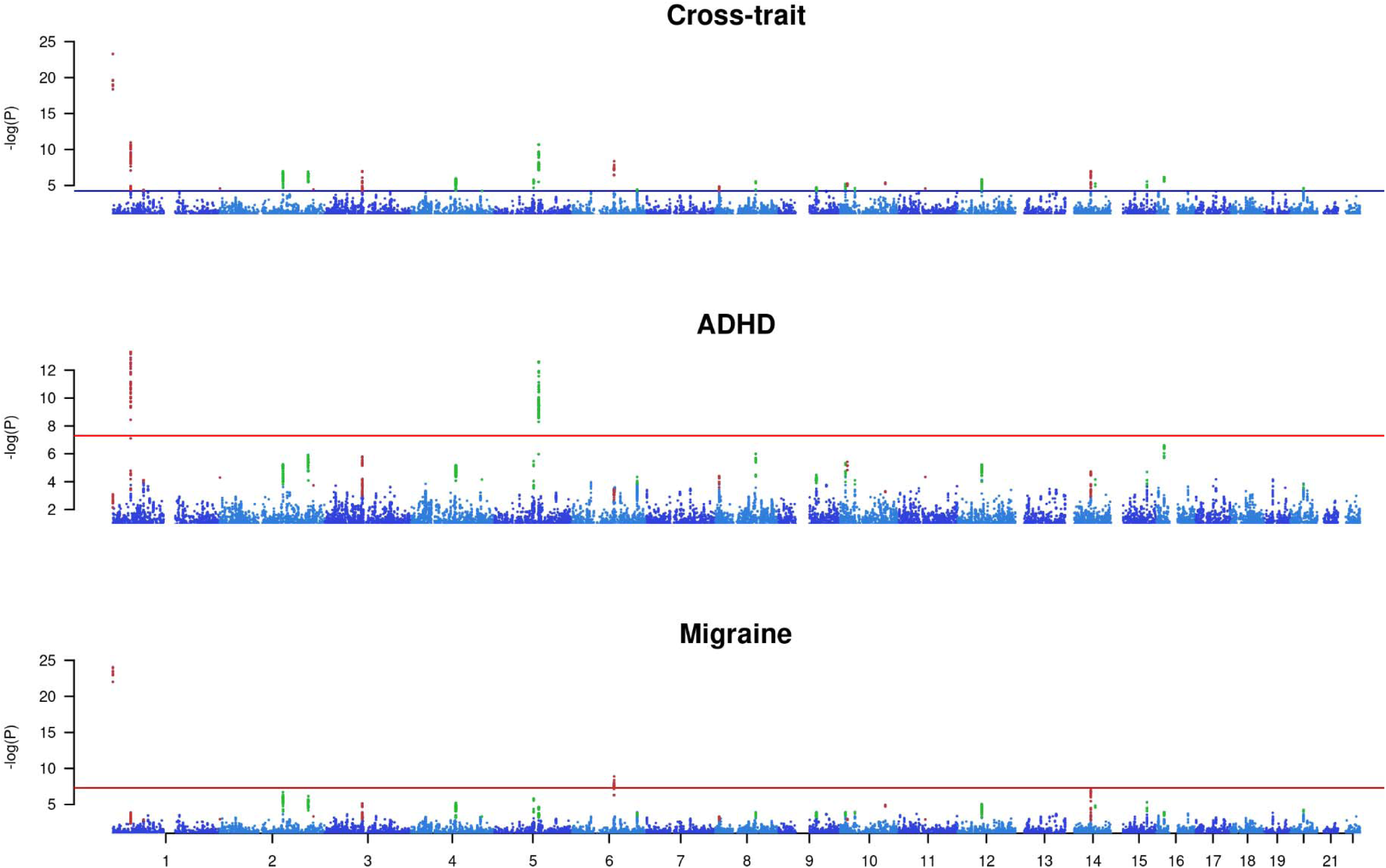
Manhattan plot for Cross-trait analysis, ADHD and migraine. Each dot represents a pleiotropic variant (theta.q-value<0.05) present in the ADHD and migraine studies, across th autosomes. Variants are represented in function of its associated -log10(P-value), for the cross-trait study -log10 (r.p-value) is represented. The blue horizontal line in the Cross-trait refers to r.q-v 0.05 translated to the equivalent P-value and the red line in ADHD and migraine refers to the genome-wide threshold (P-value = 5e-8). The two tons of blue used on the variants are meant to disting between chromosomes, green coloured variants are significant concordant SNPs on the cross-trait and red coloured variants are significant discordant variants on the cross-trait.

**Figure 2.**
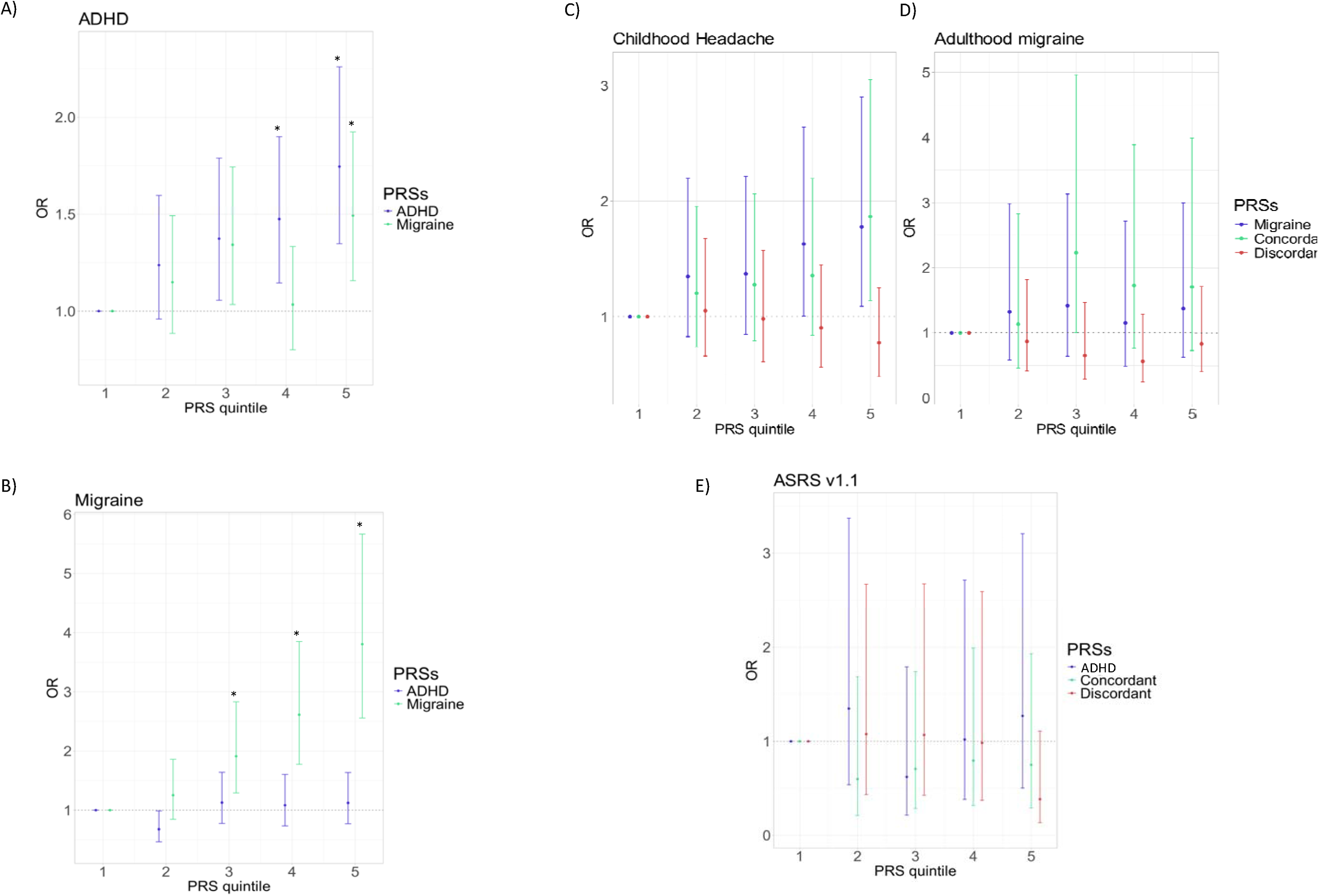
Quintiles plots of odds ratios (OR) with 95% confidence intervals for polygenic risk scores. Target sample is divided into quantiles and PRS of each quantile is compared to the first uantile using logis models with age, sex and 10 principal genetic components as fixed effects. Asterisks (*) identify statistically significant quintiles. Each plot is tested on a different target sample: **A)** ADHD case-control sample: 1,7 2,558 controls; **B)** Case only ADHD sample; Childhood headache: 311 cases and 454 controls; **C)** Case only ADHD sample; Adulthood migraine: 93 cases and 410 controls; **D)** Migraine case-contr l sample: 1,325 ca controls; **E)** Case only migraine sample; ASRSv1.1: 73 positive and 183 negative screening.

In the migraine case-control clinical sample, PRS_Mig_ was strongly associated with migraine diagnosis (OR=1.71;95%CI=(1.51, 1.94);P=2.29e-17), while PRS_ADHD_ was not significantly associated, although the effect was in the expected direction (OR=1.07; 95%CI=(0.95,1.20); P=0.276; *Figure 2B*).

### PRS_MIG_ association with clinical heterogeneity in the ADHD sample

Within ADHD cases, the PRS_Mig_ was associated with increased odds of childhood headache (n=765;OR=1.19;95%CI=(1.03, 1.39);P=0.022), and showed a non-significant trend towards increased odds of adult migraine (n=507;OR=1.18;95%;CI=(0.95, 1.49);P=0.15; *Table 2)*.

The PRS constructed with the concordant set of pleiotropic and migraine-specific variants (PRS_Mig_CC_) showed a consistent and stronger association with childhood headache (OR=1.23;95%CI=(1.07,1.43);P=4.68E-03) and a slight improvement of the strength of association with adulthood migraine compared to PRS_Mig_, although not reaching statistical significance (OR=1.24;95%CI=(0.99,1.57);P=0.067; *Supplementary* T*able 7, Figure 2, Supplementary Figure 5; Supplementary Table 8)*. In contrast, the PRS constructed for the discordant set (PRS_Mig_DC_) showed no association with either childhood headache nor adult migraine (*Supplementary Table 7; Figure 2C – 2D; Supplementary Figure 5; Supplementary Table 8)*.

When we divided the ADHD sample according to the presence of childhood headache (N=773; 40.9% reported presence of childhood headache), we found evidence of worse cognitive function, and higher levels of anxiety, depression and neuroticism in individuals with ADHD and childhood headache (*Supplementary* T*able 9).* None of these traits, however, were associated to the PRS_Mig_ and PRS_Mig_CC_ (*Figure 2E; Supplementary Table 8; Supplementary Table 10)*.

### PRS_ADHD_ association with clinical heterogeneity in the migraine sample

None of the PRSs for ADHD (PRS_ADHD_, PRS_ADHD_CC_ or PRS_ADHD_DC_) were associated with the disorder in the migraine sample screened for ADHD (N=256; 29% with a positive response) (OR=1.04;95%CI=(0.79,1.38);P=0.76, *Supplementary Table 7*).

## 4. Discussion

Migraine is a complex neurological disorder that is up to 1.8 times more frequent in individuals with ADHD than in the general population (Hansen et al., 2018). Although a significant genetic correlation between the two conditions has been reported, their shared genetic background and molecular mechanisms remain largely unexplored (Anttila et al., 2020; Ciochetti et al., 2025). In this study, we identified 29 loci associated with both disorders, including 9 novel loci not previously associated with either condition. Furthermore, in our in-house ADHD sample, we found that migraine genetic liability was associated with childhood headache, an association apparently driven by a subset of migraine-specific and migraine-ADHD pleiotropic variants with concordant effects. Notably, individuals with ADHD and childhood headaches exhibited higher levels of anxiety, depression, and neuroticism, along with poorer cognitive functions compared to those with ADHD but without childhood headaches.

To better understand the pleiotropic molecular mechanisms underlying both traits, we performed functional annotation of the 29 cross-trait loci, stratifying them by their direction of effect in ADHD and migraine. Genetic variants with concordant direction of effect mapped to 83 genes, which were enriched in several immune system pathways, mainly driven by RAET1/ULBP genes superfamilies related with the Major Histocompatibility Complex Class I (MHCI), a key immune system protein complex that also plays important roles in neural development and synaptic plasticity (Cebrin et al., 2014; Elmer & McAllister, 2012). The role of MHCI in synaptic function has been extensively studied in the visual system, hippocampus, and cerebellum, with reported contributions to memory, motor function, and anxiety-related behaviors (Nelson et al., 2013; Sankar et al., 2012; Shatz, 2009). Deficient MHCI expression in the brain induces locomotor hyperactivity, motor impulsivity, and attention deficits in mice, three major symptoms of ADHD (Meng et al., 2021). Specific alleles of this protein (MHC I) have been associated to clinical-based migraine in a Taiwanese sample (Huang et al., 2020). In line with this, we also observed enrichment in other autoimmune and inflammatory conditions such as vitiligo, asthma or chronic obstructive pulmonary diseases (COPD). This broader autoimmune/atopic enrichment, together with HLA/MHCI signals, is compatible with neuroimmune models of migraine, including meningeal immune-cell activation and cytokine signalling implicated in attack initiation and chronification (Edvinsson et al., 2019). We also found enrichment for refractive errors, such as hypermetropia and astigmatism, which have previously been associated with ADHD (Bellato et al., 2023). Interestingly, individuals with these conditions appear to be more prone to headaches (Wajuihian, 2024). We hypothesize that uncorrected hyperopia and astigmatism may lead to visual fatigue, thereby increasing headache risk and impairing attention, potentially exacerbating ADHD symptoms. Additionally, we observed enrichment in several gene sets related to brain structure. Both ADHD and migraine have been associated with alterations in cortical thickness: reduced prefrontal volume correlates with inattention and impulsivity symptoms in individuals with ADHD (Valera et al., 2007), while decreased gray matter volume correlates with pain severity and attack frequency in migraine individuals (Hubbard et al., 2014). Finally, we observed enrichment in genes related to cerebrovascular disorders. Migraine, particularly with aura, has been consistently associated with an increased risk of ischemic stroke (Øie et al., 2020) , whereas ADHD has been linked to higher incidence of cardiovascular disease, including haemorrhagic stroke episodes (Li et al., 2022).

On the other hand, genetic variants with discordant direction of effect mapped 48 genes, which were enriched in several mental health disorders and related conditions, including autism spectrum disorder, schizophrenia, bipolar disorder, neuroticism, anorexia nervosa, and cannabis use. These traits have been extensively associated with ADHD, as supported by a substantial body of literature (Antshel & Russo, 2019; Canals et al., 2024; Froude et al., 2024; Schiweck et al., 2021). A similar pattern was observed for gene set related to lifestyle and behavioural factors, which also show strong evidence of association with ADHD (Cortese et al., 2016). In contrast, the relationship between migraine and these traits is less well explored. Although some reviews suggest potential associations, they also highlight the need for further research to better understand these links (Łangowska-Grodzka et al., 2023). In addition, we found enrichment in several gene-sets related to cardiovascular and blood pressure traits, mainly involving hypertension. The relationship between migraine and blood pressure remains a matter of debate, with mixed results, although patients with migraine report fewer episodes when treated with antihypertensive medications (Kalala et al., 2025).

At the gene level, we identified several pleiotropic genes encoding proteins essential for proper brain function. Among the 83 genes mapped with concordant associations between ADHD and migraine, we found *CAMK1D*, a member of the calcium/calmodulin-dependent protein kinase 1 family, involved in dendritic growth from hippocampal neurons and previously related to ADHD (Wei et al., 2022). We also highlight *PARD3*, which encodes PAR3, a protein involved in exon splicing and neuronal polarity, that plays an important role in learning and memory, and has been previously associated with educational attainment, high mathematical ability, and brain morphology (Lee et al., 2018; Okbay et al., 2022; Shi et al., 2003; van der Meer et al., 2020; Voglewede et al., 2024). Additionally, we identified three intronic variants with colocalization evidence supporting a shared signal between ADHD and migraine, which are also associated with gene expression in brain for *SLC9B1*, *ERBB3* and *ABHD17C. SLC9B1* encodes a sodium/hydrogen exchanger involved in ion transport, which has been implicated in the pathophysiology of migraine (Spekker et al., 2023), as well as in its shared genetic risk with schizophrenia (Bahrami et al., 2022). *ERBB3* encodes a tyrosine-protein kinase that acts as a key cell surface receptor for neuregulins, including neuregulin-1 (NRG1), which has been strongly linked to schizophrenia and other neuropsychiatric conditions (Douet et al., 2014). NRG/ERBB signaling has been shown to regulate glial cells (Mei & Nave, 2014) which are essential for nervous system development and have been hypothesized to contribute to migraine susceptibility (Vila-Pueyo et al., 2023). *ABHD17C* encodes a palmitoyl-protein hydrolase involved in synaptic regulation (Sohn & Park, 2019) and has been associated with the shared genetics between ADHD and lifespan (Vilar-Ribó et al., 2023). Taken together, these findings highlight a number of genes as biologically plausible candidate genes involved in neuronal development, synaptic function, and cognitive regulation, reinforcing the hypothesis of shared neurobiological underpinnings between ADHD and migraine.

Among the 48 genes mapped by discordant variants, we found the *ARID4A* gene, which encodes a DNA-binding protein that modulates the activity of several transcription factors, proposed as a potential candidate gene for ADHD in a recently published epigenome-wide association study (EWAS), and has also been associated with schizophrenia and alcohol consumption (M. Liu et al., 2019; Meijer et al., 2023; Ren et al., 2022). *CADM2*, member of the synaptic cell adhesion molecule 1 (SynCAM) family, showed consistent evidence of pleiotropy and colocalization, and it has been previously associated with impulsive personality traits and also to migraine confirming its pleiotropy role on both conditions(Sanchez-Roige et al., 2019, 2023).

Polygenic risk score analyses revealed a trend toward higher migraine genetic load in individuals with ADHD compared to controls. Those in the highest migraine PRS quintile had significantly higher odds of ADHD than those in the lowest, consistent with epidemiological evidence of increased migraine prevalence in ADHD (Arruda & Arruda, 2014; Fasmer et al., 2011; Jameson et al., 2016; Kerem, 2016). Within the ADHD sample, migraine PRS was associated with childhood headaches. Individuals with ADHD and childhood headaches showed higher levels of anxiety, depression, and neuroticism than those without headaches. Previous studies have linked ADHD, depression, and migraine (Jahangir et al., 2020), reporting that up to 80% of individuals with migraine experience depression during their lifetime (Leo & Singh, 2016), and that depression is 5.5 times more prevalent among youths with ADHD (BIEDERMAN et al., 1996). Individuals with both migraine and comorbid depression or anxiety also tend to exhibit higher neuroticism (Galvez-Sánchez & Montoro Aguilar, 2022). Evidence further suggests shared brain networks—particularly involving hypothalamic structures and dopaminergic and serotonergic systems related to sleep regulation—may underlie the pathophysiology of both ADHD and headache (Paolino et al., 2015), systems that are also closely linked to depression and its physiopathology (Jauhar et al., 2023; Pizzagalli et al., 2019).

Our study demonstrates that separating pleiotropic variants into concordant and discordant subsets is a useful strategy to reduce variant complexity and enhance interpretability. This approach was supported by colocalization analyses, which showed that 61% concordant lead variants showed evidence of pleiotropy, compared with only 16% of discordant variants. Using concordant migraine-specific and pleiotropic variants in migraine PRS analyses strengthened the association compared with PRS constructed from all variants, while requiring only about one quarter of the variants. These findings support the utility of the concordant-discordant PRSs strategy, which preserves predictive power while requiring substantially fewer genetic variants. Overall, our results, supported by colocalization, PRS and gene-set enrichment analyses, indicate that ADHD and migraine predominantly share concordant biological pathways, further reinforcing the positive genetic correlation already reported (Ciochetti et al., 2025).

Our results should be interpreted with several considerations: (i) The present study uses self-reported headache and migraine information in ADHD cases, rather than migraine diagnosis. It might be that the modest association that we found between migraine genetic load and childhood headache/ adult migraine in our ADHD cases, is due to a noisy definition of the phenotype and a limited samples size, and that a more precise clinical characterization of migraine in a larger study would strengthen this association. A similar limitation applies to the migraine clinical sample screened with ASRS1.1 for ADHD. (ii) Despite detecting an association between PRS_Mig_ and migraine, the precision of our estimates may have been influenced by potential misclassification, given that the control sample did not exclude possible migraine cases. (iii) Variants in the original GWAS-MA for ADHD and migraine differ, causing a loss of SNPs in the cross-trait analysis and therefore in the concordant and discordant sets for the PRSs. (iv) Although a trend toward a higher migraine genetic load in ADHD cases compared to controls is observed, this difference was significant only in the highest PRS quintile. This may indicate that the effect size of the migraine genetic load (PRS_Mig_) on ADHD cases is small and our study lacked sufficient statistical power to detect broader association. Despite our results from PRSs support a common genetic background between both disorders, further studies with larger sample sizes are needed to clarify this relationship. (v) Although previous studies have reported sex-specific differences in the co-occurrence of ADHD and migraine (Arruda et al., 2020; Carpenet et al., 2019), the lack of sex-stratified GWAS results available for ADHD and migraine have prevented us from undertaking sex-specific analyses. Further studies are required to conduct sex-stratified GWAS in order to elucidate potential sex-specific genetic mechanisms underlying the comorbidity between ADHD and migraine.

To summarize, our study identifies 29 loci influencing both ADHD and migraine, including 9 novel loci not previously associated with either disorder. Some of these loci map to coding genes with relevant brain molecular functions such as *SLC9B1*, *ERBB3* and *ABHD17C*, representing promising candidates for further research. We also found that individuals with a high genetic burden for migraine, have higher odds for ADHD compared to those with a low genetic load for migraine. In addition, we found that ADHD patients with a history of childhood headache exhibit an increased genetic load for migraine, along with elevated levels of anxiety, neuroticism and depression, as well as reduced cognitive function. Finally, our results highlight the utility of separating genetic variants according to their direction of effect to provide insights into the shared genetic architecture of comorbid disorders, with potential to improve understanding of clinical heterogeneity and enhance risk prediction.

## Data Availability

All data produced in the present study are available upon reasonable request to the authors

## Financial Support

This work was supported by the Agència de Gestió d’Ajuts Universitaris i de Recerca (AGAUR, 2017SGR-1461, 2021SGR-00840, 2014SGR-0932, 2009SGR-0078), the Instituto de Salud Carlos III (PI10/00876, PI20/00041, PI22/00464, PI23/00404, PI23/00026, PI24/00195 and CP22/00128 to M.S.A and CP22/00026 to S.A), the Network Center for Biomedical Research (CIBER) to J.C.D. and U.Z.A; the European Regional Development Fund (ERDF); the ECNP Network ‘ADHD across the Lifespan’, the “Fundació la Marató de TV3” (202228-30, 202228-31 and 072310), Spanish Ministry of Economy and Competitiveness (SAF2009–13182-C01, SAF2009–13182-C03).

## Acknowledgements

The authors are grateful to patients and controls who kindly participated in this research. The genotyping service was carried out at the Genotyping Unit-CEGEN in the Spanish National Cancer Research Centre (CNIO), supported by Instituto de Salud Carlos III (ISCIII), Ministerio de Ciencia e Innovación. CEGEN is part of the initiative IMPaCTGENóMICA (IMP/00009) cofunded by ISCIII and the European Regional Development Fund (ERDF).

## Competing Interests

P.P.R. has received, in the last three years, honoraria as a consultant and speaker for: AbbVie, Dr. Reddy’s, Eli Lilly, Lundbeck, Medscape, Novartis, Pezer, Organon and Teva. Her research group has received research grants from AbbVie, Novartis and Teva; as well as, Instituto Salud Carlos III, EraNet Neuron, European Regional Development Fund (001-P-001682) under the framework of the FEDER Operative Programme for Catalunya 2014–2020 - RIS3CAT; has received funding for clinical trials from AbbVie, Amgen, Biohaven, Eli Lilly, Novartis, Teva. She is the Honorary Secretary of the International Headache Society. She is in the editorial board of Revista de Neurologia. She is an associate editor for Cephalalgia, Headache and Neurologia. She is a member of the Clinical Trials Guidelines Committee of the International Headache Society. She has edited the Guidelines for the Diagnosis and Treatment of Headache of the Spanish Neurological Society. She is the founder of www.midolordecabeza.org. P.P.R. does not own stocks from any pharmaceutical company. C.F. declares that he has given lectures or received help to attend conferences from Rubió, Exceltis and Takeda. J.A.R.Q was on the speakers’ bureau and/or acted as consultant for Biogen, Idorsia, Casen-Recordati, Johnson&Johnson, Novartis, Takeda, Bial, Sincrolab, Neuraxpharm, Novartis, BMS, Medice, Rubió, Uriach, Technofarma and Raffo in the last 3 years. He also received travel awards (air tickets + hotel) for taking part in psychiatric meetings from Idorsia, Johnson&Johnson, Rubió, Takeda, Bial and Medice. The Department of Psychiatry chaired by him received unrestricted educational and research support from the following companies in the last 3 years: Exeltis, Idorsia, Casen-Recordati, Takeda, Neuraxpharm, Oryzon, Roche, Probitas and Rubió. Johnson&Johnson. M.R. received travel awards (air tickets + hotel) for taking part in psychiatric meetings from Rubió. All other authors declare no biomedical financial interests or conflicts of interest.

## Authorship and contributorship

P.C.G. conducted the literature search. P.C.G., N.L. undertook the statistical analysis. V.M.C., P.C.G., M.D.R.R., U.Z.A. processed the biological samples. J.A.R.Q, C.F, M.C., V. R., V.G, P.P.R carried out the sample recruitment and clinical evaluation. P.C.G., J.C.D., M.S.A. wrote the first draft of the manuscript. M.R, S.A. contributed to the writing. J.C.D, M.S.A. designed the study and supervised the work. All authors reviewed and approved the final manuscript.

## Notes

### Funding Statement

This work was supported by the Agencia de Gestio dAjuts Universitaris i de Recerca
(AGAUR, 2017SGR-1461, 2021SGR-00840, 2014SGR-0932, 2009SGR-0078), the Instituto
de Salud Carlos III (PI10/00876, PI20/00041, PI22/00464, PI23/00404, PI23/00026,
PI24/00195 and CP22/00128 to M.S.A and CP22/00026 to S.A), the Network Center for
Biomedical Research (CIBER) to J.C.D. and U.Z.A; the European Regional Development
Fund (ERDF); the ECNP Network ADHD across the Lifespan, the Fundacio la Marato de
TV3 (202228-30, 202228-31 and 072310), Spanish Ministry of Economy and
Competitiveness (SAF2009-13182-C01, SAF2009-13182-C03).

### Author Declarations

Ethics committee/IRB of Clinical Research Ethics Committee (CREC) of Hospital Universitari Vall d Hebron ethical approval for this work

