## Supplementary material for "Exploring the genetic overlap between attention-deficit/hyperactivity disorder (ADHD) and migraine": Supplemantary materials

Supplementary Contents

### Supplementary Methods

#### SNP quality control filtering in GWAS-MA samples

##### GWAS-MA data processing

SNPs that were non-biallelic, duplicated, or with strand-ambiguous alleles were removed. We also filtered out SNPs with INFO scores < 0.8, minor allele frequency (MAF) < 0.01 or non- autosomal. In addition, SNPs with effective sample size $\frac{4N_{cases}N_{contols}}{N_{cases}+N_{controls}}$ <70% of the total sample size were excluded from ADHD summary statistics. In the migraine dataset, SNPs with effective sample size, calculated as $1/{f \left( 1-f \right){se}^{2}}$, being $f$ effect allele frequency and $se$ the standard error) < 5,000 were also removed, as it was done in the original migraine GWAS-MA (Hautakangas et al., 2022).

#### PolarMorphism

##### SNP classification

SNPs with effects specific to migraine will have $\theta$ values close to *0* or $\pi$ and SNPs that are specific to ADHD will have $\theta$ values close to $\frac{\pi}{2}$ or – $\frac{\pi}{2}$ . Pleiotropic SNPs were classified as concordant (CC, same direction of effect in both traits), which will be close to the diagonal line with slope of 1 that goes through the origin, that is with $\theta$ values around $\frac{\pi}{4}$ or - $\frac{3\pi}{4}$ and discordant (DC, opposite direction of effect between traits), which will be close to the diagonal line of slope -1 that goes through the origin, that is with $\theta$ values around $-\frac{\pi}{4}$ or $\frac{3\pi}{4}$ .

##### SNP clumping and new loci

Top hits from PolarMorphism were clumped (r^2^>0.1, Kb ±500) using the 1000 Genomes Project Phase 3 European as the reference panel for linkage disequilibrium (LD) computation and PLINK 1.09 (Auton et al., 2015; Purcell et al., 2007) to detect independent loci. We considered a novel locus when any of the SNPs in LD with the lead SNP (r^2^>0.1) within a 500kB window were not genome-wide significant (p<5e-08) in any of the original GWAS-MA summary statistics (Demontis et al., 2023; Hautakangas et al., 2022). The same procedure was done for the lead SNPs reported in the recent cross-trait analysis between ADHD and migraine (Ciochetti et. al. 2025).

##### Functional annotation

Functional annotation was performed for significant SNPs ($r$ q-value < 0.05 and $\theta$ q-value < 0.05) and SNPs in the 1000 Genomes Project Phase 3 European panel (Auton et al., 2015) in LD (r2 > 0.6) with any of the significant variants. The analysis was run in FUMA (Functional Mapping and Annotation of Genome-Wide Association Studies, <https://fuma.ctglab.nl/>)(Watanabe et al., 2017). A lead SNP is the one with the most significant p-value within a clump of SNPs with r2>0.1 in a 500kb window. Two lead SNPs within 250kb are considered to be in the same locus. All the analyses were run for concordant and discordant variants subsets separately. We integrated data from: (i) Combined Annotation Dependent Depletion (CADD) scores (Kircher et al., 2014), which predict how deleterious a SNP effect is in protein structure/function based on 63 functional annotations, (ii) Regulome DB scores(Boyle et al., 2012a), a categorical score that estimates the regulatory functionality of SNPs based on existing functional data (annotation to cis-eQTLs, expression quantitative trait loci) and evidence for transcription factor binding, and (iii) chromatin states, which predicts the accessibility of genomic regions using 15 categorical states, predicted by ChromHMM based on 5 chromatin marks for 127 epigenomes. CADD ≥ 12.37 was considered as threshold for deleterious variants, RegulomeDB score < 3 were considered to have a regulatory function, and minimum chromatin states between 1-7 were considered open chromatin states. We also used FUMA to map SNPs to genes based on physical proximity (using default parameters), eQTL in brain (based on GTEx v8 Brain, CommonMind Consortium and BRAINEAC) and chromatin interaction mapping for brain specific regions (data bases used detailed in Supplementary material). We enquired for known brain eQTLs using the 13 brain areas from GenotypeTissue Expression (GTEx) v8 (Battle et al., 2017) and BRAINEAC(Ramasamy et al., 2014). Functionally annotated variants were mapped to genes, then tested for enrichment in biological pathways using gene ontology (GO) biological process, Kyoto Encyclopedia of Genes and Genomes (KEGG), BioCarta and the 20^th^ of July 2023 of the NHGRI-EBI GWAS catalogue(MacArthur et al., 2017). All analyses were corrected for multiple comparisons using 5% False Discovery Rate (FDR). All the analyses were run for concordant and discordant variants subsets separately.

With regards Euchromatin interaction mapping, a total of 7 FUMA Hi-C build in data sets are used, specifically: PsychENCODE EP links and promoter anchor loops, HiC Giusi-Rodrigez et. al. 2019 for adult and fetal cortex, and HiC (GSE87112) for dorsolateral prefrontal cortex, hippocampus and neural progenitor cells. An FDR p-value threshold of 1E-10 was used. The promoting regions analysed were 250 kb upstream of the transcriptomic start sides (TSS) and 500 kb downstream. By the chromatin interaction mapping, genes whose user defined promoter regions are overlapped with the significantly interacting regions will be mapped. Predicted enhances and promoters were annotated using 10 brain regions from Roadmap epigenomics project (cortex, ganglion eminence, angular gyrus, anterior caudate, cingulate gyrus, germinal matrix, hippocampus middle, inferior temporal lobe, dorsolateral prefrontal cortex and subtantia nigra) and for fetal male and female brain. Then SNPs were filtered by enhancers and genes by promoters.

More info in: <https://fuma.ctglab.nl/tutorial#chromatin-interactions>

##### Colocalization analysis

To investigate whether the same causal variant underlies the association of the pleiotropic lead variants in both ADHD and migraine, we used the Coloc R package (Giambartolomei et al., 2014). Information for SNPs within 500kb windows centred in each pleiotropic lead SNP was retrieved from the original ADHD and migraine GWAS-MA. This method uses Bayesian statistical models to assess whether two association signals are consistent with a shared causal variant (Giambartolomei et al., 2014). The method estimates posterior probabilities for five different hypotheses: (i) H­_0_, no association with either trait; (ii) H_1_, association with ADHD but no with migraine; (iii) H_2_, association with migraine but no with ADHD; (iv) H_3_, two independent association signals, one for each condition; or (v) H_4_, a shared association signal. A posterior probability (PP) threshold of >0.6 for H_3_ or H_4_ was used to determine pleiotropy (Han et al., 2023). Graphic interpretation was performed with LocusCompareR R package (Liu et al., 2019), which plots ${-log}_{10}(p-value)$ ADHD vs ${-log}_{10}(p-value)$ migraine and a region plot for each trait.

#### Clinical assessment

##### ADHD

The ADHD clinical evaluation consisted of: (i) ADHD diagnosis based on symptomatology using the Conner’s Adult ADHD Diagnostic Interview for DMS-IV (CAADID), (ii) severity of ADHD symptoms, the levels of impairment and the presences comorbid disorders to increase diagnose accuracy (the ADHD Rating Scale (ADHD-RS), the Sheehan Disability Inventory (SDI), Functioning Assessment Short Test (FAST), and the Structured Clinical Interview for DSM-IV Axis I and II Disorders (SCID-I and SCID-II) and (iii) a group of self-administrated interviews to test ADHD symptoms (Conners’ ADHD Rating Scale (CAARS)), depressive symptoms (Beck Depression Inventory (BDI)(Beck, 1996)), anxiety traits (State-Trait Anxiety Inventory (STAI-R)), impulsivity (Barratt Impulsiveness Scale (BIS-11)), as well as the continuous performance tasks (CPTs) used to measure individual differences in sustained attention, and the Zuckerman–Kuhlman Personality Questionnaire (ZKPQ) to assess personality features including neuroticism-anxiety, activity, sociability, impulsive sensation-seeking, and aggression-hostility.

Exclusion criteria of ADHD cases were IQ < 70; a history or the current presence of a condition or illness, including neurologic, metabolic, cardiac, liver, kidney, or respiratory disease; a chronic medication of any kind; birth weight ≤ 1.5 kg; and other neurological or systemic disorders that might explain ADHD symptoms.

##### Migraine

To ensure diagnostic accuracy, individuals with migraine were required to maintain a daily headache diary for at least one month prior to blood sample collection. All participants underwent a thorough clinical evaluation by a headache specialist, with clinical outcomes collected as previously described (Torres-Ferrús et al., 2017). These included detailed demographic information, medical history, comorbidities, pain characteristics, migraine triggers, and treatments. Additionally, all participants completed the 36-item Short Form Health Survey (SF-36 v2)(J E Ware Jr & C D Sherbourne, 1992), the State-Trait Anxiety Inventory (STAI)(Spielberger, 1971), and the Beck Depression Inventory II (BDI-II) (Beck, 1996) to assess baseline quality of life, anxiety levels, and the presence of depressive symptoms, respectively. For participants with migraine, additional assessments included the Migraine Disability Assessment (MIDAS) (Stewart et al., 2001) and the Headache Impact Test 6 (HIT-6)(Yang et al., 2011) to evaluate the impact and disability associated with the condition.

##### Controls

Individuals with ADHD symptomatology were excluded retrospectively from the control sample under the following criteria: (1) diagnosed with ADHD previously and (2) answering positively to the life-time presence of the following ADHD symptoms: (a) often has trouble in keeping attention on tasks, (b) usually loses things needed for tasks, (c) often fidgets with hands or feet or squirms in seat, and (d) often gets up from seat when remaining in seat is expected. Controls were not screened for migraine.

#### Genotyping and imputation

For both the ADHD cases and all the controls used in this study, genomic DNA was isolated from whole blood using salting-out protocol and genotyped in seven different genotyping waves using five different chips: 569 individuals with Omni1.1M, 256 individuals with Omni2.5M, 304 individuals with Illumina Infinium Psy1Chip v1.0 array, 526 individuals with Infinium™ Global Screening Array-24 v2.0 and 120 individuals with Infinium™ Global Screening Array-24 v3.0.

For the migraine cases, genomic DNA was extracted from peripheral blood leukocytes according to standard protocols (Freilinger et al., 2012) and genotyping was performed on InfiniumCoreExome-24v1-1_A beadchip.

Pre-imputation quality control was applied for all samples considering each GWAS batch separately using PLINK 2.0 (Chang et al., 2015), following standards protocols as previously described (Cabana-Domínguez et al., 2024). In brief, individual and variant filtering was performed according the following criteria: SNP call rate > 0.95 prior sample filtering, individual call rate > 0.98, autosomal heterozygosity deviation within ± 0.2, variant call rate > 0.98 after sample filtering, Hardy-Weinberg equilibrium (HWE) P > 1e-06 in controls and P > 1e-10 in cases, and minor allele frequency (MAF) > 0.01. Individuals showing sex discrepancy were also excluded. Population structure was examined using principal component analysis (PCA) in PLINK 2.0 and the mixed ancestry 1000G reference panel (Auton et al., 2015). Ancestry outliers were removed if their principal component (PC) values for PC1 or PC2 deviated by more than one standard deviation from the mean-centering point of the study sample, considering each GWAS wave separately. Relatedness and potential sample duplication were assessed by the “KING-robust kinship estimator” analysis in PLINK 2.0 (Manichaikul et al., 2010), excluding one individual from each pair of subjects with kinship coefficient > 0.0442. Post-imputation dosage files were filtered with imputation SNP call rate > 0.95, INFO score > 0.8, MAF > 0.01 and case-control differential missingness rate < 0.2 in the overall sample.

All three samples were prepared for imputation using McCarthy tools, and imputed with the Michigan Imputation Server (Das et al., 2016), using Haplotype Reference Consortium (HRC Version r1.1 2016) reference panel GRCh37/hg19. Pre and post-imputation quality control can be found on the supplementary methods.

#### Construction of polygenic risk scores

Polygenic risk scores for ADHD (PRS_ADHD_) and migraine (PRS_Mig_) were constructed using weights for SNPs included in the two GWAS-MA summary statistics described in section 2.2.1 (Demontis et al., 2023; Hautakangas et al., 2022). In addition, we constructed PRS_Mig_ combining migraine-specific and migraine-ADHD pleiotropic variants, using the overall effect ($r$) as the weight and stratifying variants according to the direction of effect into concordant and discordant subsets (PRS_Mig_CC_ and PRS_Mig_DC_, respectively). Pleiotropic variants identified by Polarmorphism with $\theta$ ranging from $0 to \frac{3\pi}{8}$ and $-\frac{5\pi}{8} to-\pi$ were selected for the PRS_Mig_CC_ and those with $\theta$ ranging from $0 to-\frac{3\pi}{8}$ and $\frac{5\pi}{8} to \pi$ were selected for the PRS_Mig_DC_ (Supplementary Figure 1B). The same procedure was followed to construct PRS_ADHD_ in individuals with migraine, combining variants with ADHD-specific and migraine-ADHD pleiotropic effects. Variants with $\theta$ ranging from $\frac{\pi}{8} to \frac{\pi}{2}$ and $-\frac{\pi}{2} to-\frac{7\pi}{8}$ were selected for the PRS_ADHD_CC_ and those with $\theta$ ranging from $-\frac{\pi}{8} to-\frac{\pi}{2}$ and $\frac{\pi}{2} to\frac{7\pi}{8}$ were selected for the PRS_ADHD_DC_ (Supplementary Figure 1C). A total of six polygenic risk scores were constructed: PRS_ADHD_, PRS_ADHD_CC_, PRS_ADHD_DC_, PRS_Mig_, PRS_Mig_CC_ and PRS_Mig_DC_. Weights were adjusted considering local LD patterns using PRS-CS (Ge et al., 2019) with phi parameter equal to 10^-2^, and PRSs were constructed in our in-house samples using PLINK 2.0 (Purcell et al., 2007). All PRSs were standardised to a mean equal to 0 and a standard deviation equal to 1.

#### PRS_Mig_ association with clinical heterogeneity in the ADHD sample

For each of the 35 clinical items previously described (Supplementary Table 1) we tested for average differences with the appropriate test depending on the data: (i) continuous data distributions with kurtosis excess and skewness ±1 were treated as normal distributions, T-Student test for equal or unequal variances were performed, using F test to compare the variances between the two groups, (ii) continuous data distributions not passing normality assessment were tested with Mann Whitney test and (iii) dichotomous variables were tested with chi-square test.

### Supplementary figures

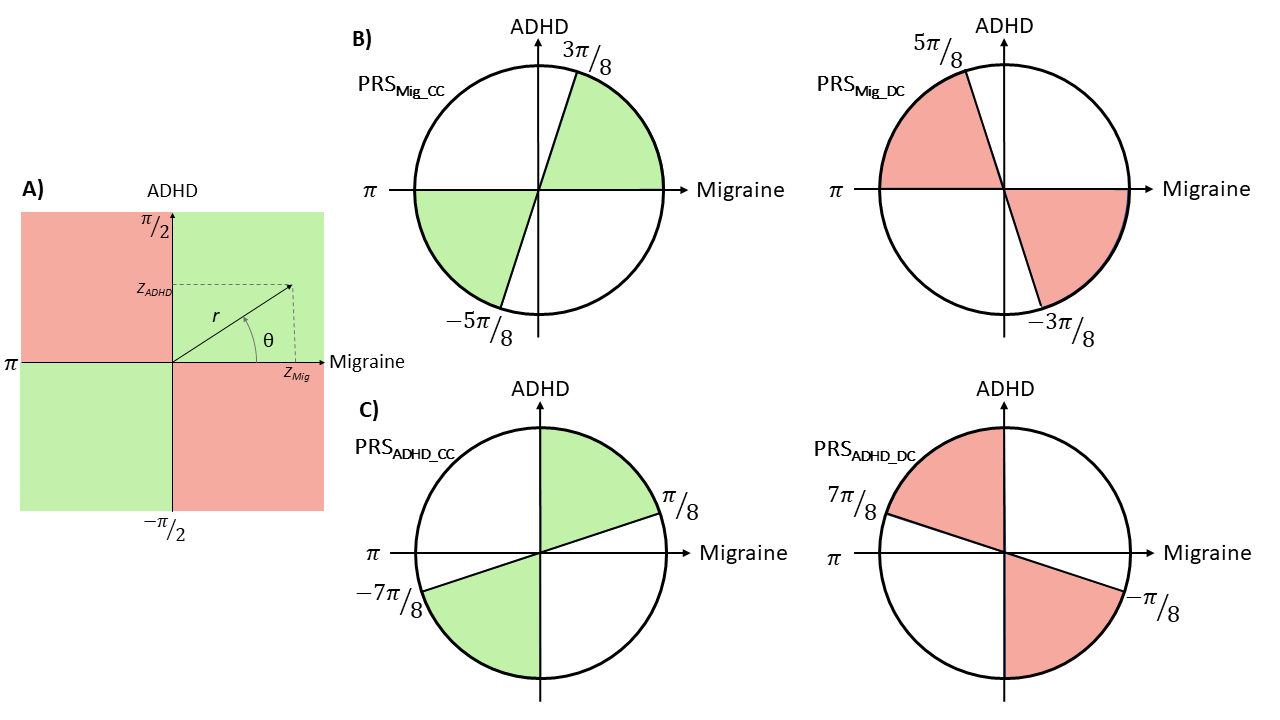

Supplementary Figure 1. Definition of variant sets based on cross-trait Z-scores from attention-deficit/hyperactivity disorder (ADHD) and migraine GWAS using PolarMorphism. **A)** Scheme illustrating conversion from Cartesian to polar coordinates, the x-axis represents migraine Z scores and the y-axis represents ADHD Z scores. SNPs laying on the green quadrants are concordant variants and those laying on red quadrants are discordant. **B)** Diagram showing the areas used to select the SNPs with an effect on migraine or a pleiotropic effect, used to construct the migraine concordant (green) and discordant (red) polygenic risk scores (PRS). **C)** Diagram showing the areas used to select the SNPs with an effect on ADHD or a pleiotropic effect, used to construct the ADHD concordant (green) and discordant (red) PRS.

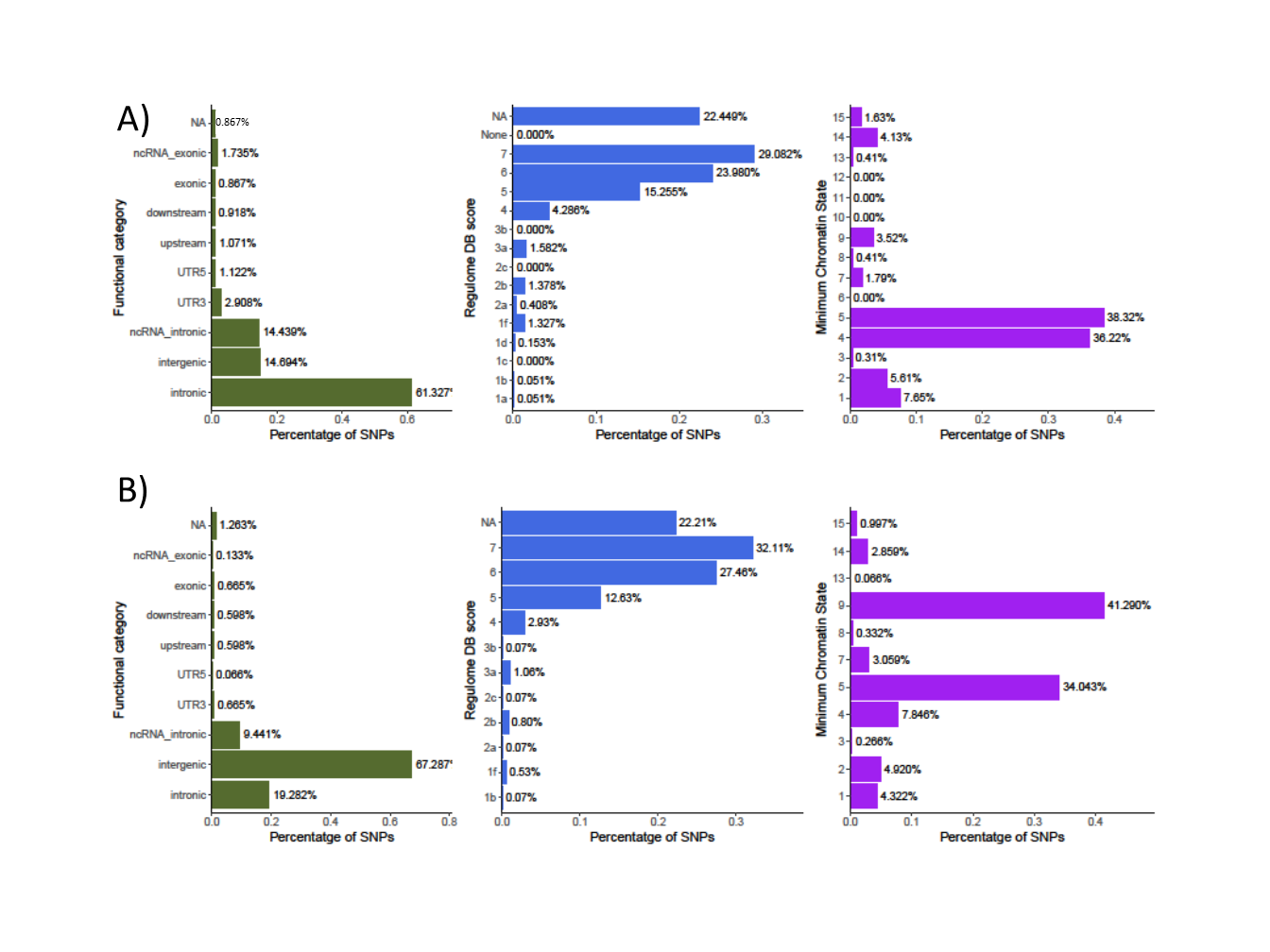

Supplementary Figure 2. Summary of functional annotation results from the cross-trait meta-analysis of attention-deficit/hyperactivity disorder (ADHD) and migraine. Proportion of **A)** concordant and **B)** discordant SNPs annotated and classified according to three categories: functional category (based on genomic location), Regulome DB score and minimum chromatin state. Regulome DB score predicts likelihood of regulatory functionality, where lower scores indicate higher likelihood. Further information can be found in Boyle et al. 2012 (Boyle et al., 2012b)(Boyle et al., 2012b)(Boyle et al., 2012b)(Boyle et al., 2012b)(Boyle et al., 2012b)(Boyle et al., 2012b)(Boyle et al., 2012b)(Boyle et al., 2012b). Minimum Chromatin State across 127 tissue and cell types, lower scores indicate higher accessibility, with states 1–7 referring to open chromatin states.

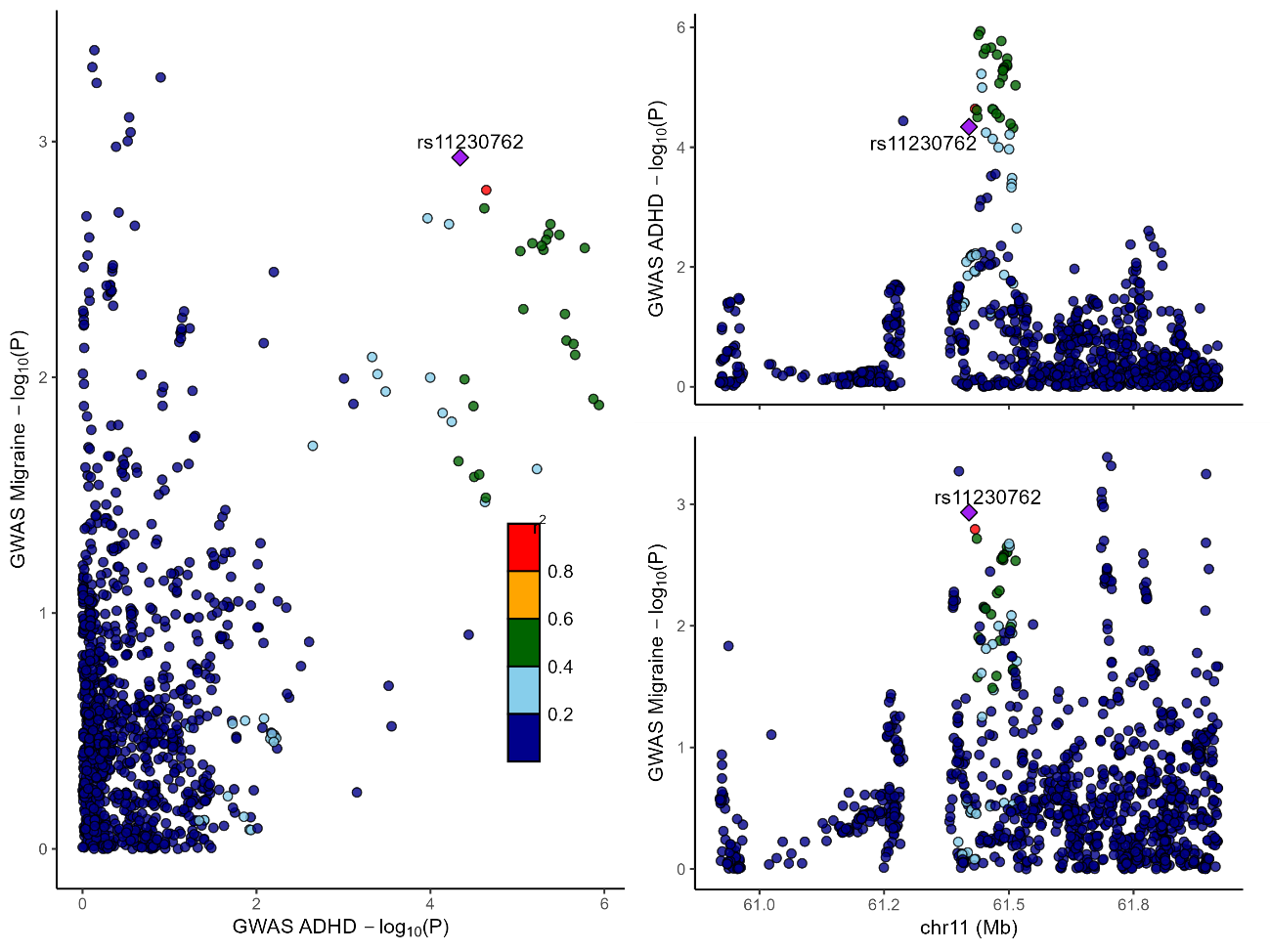

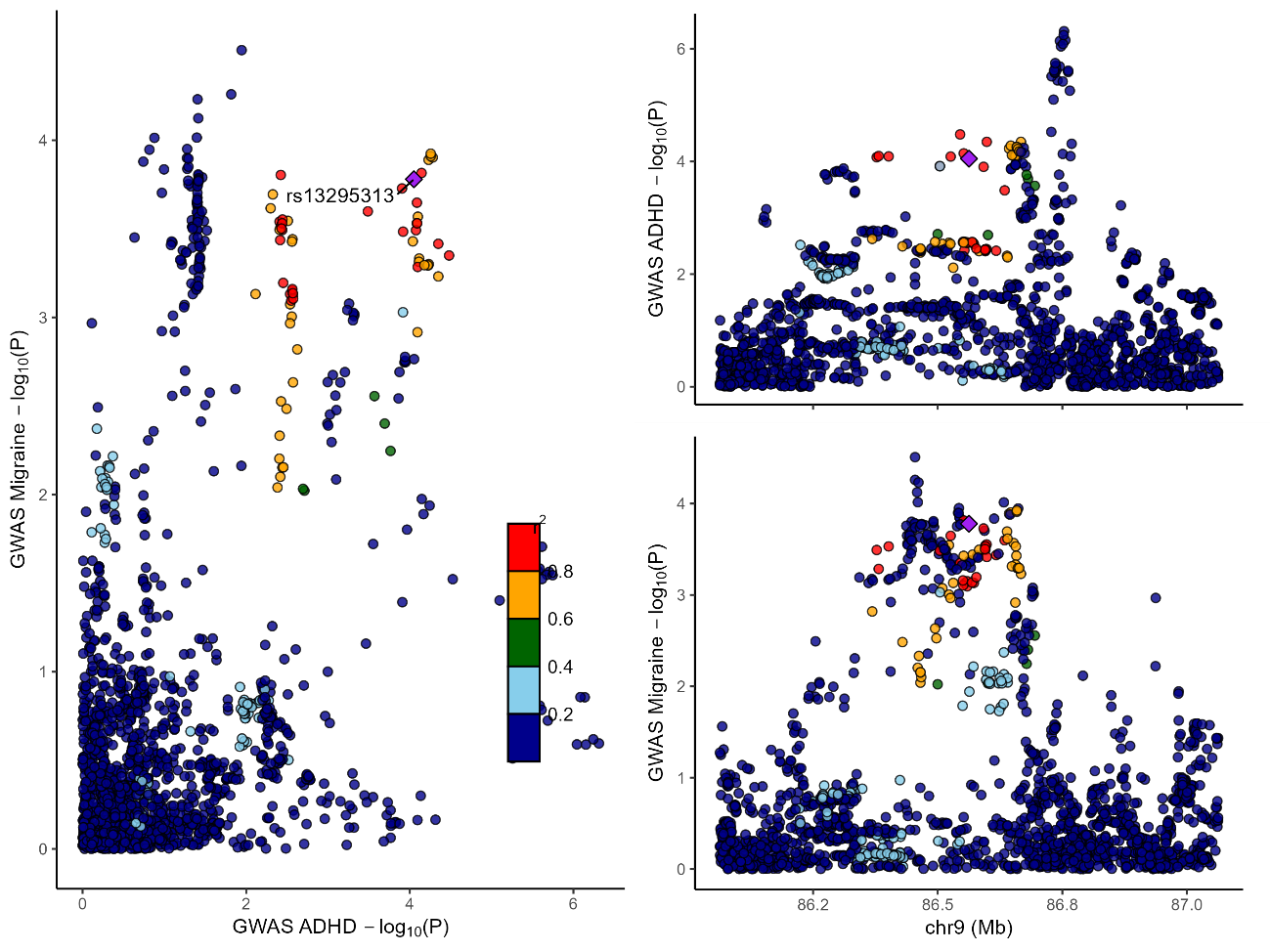

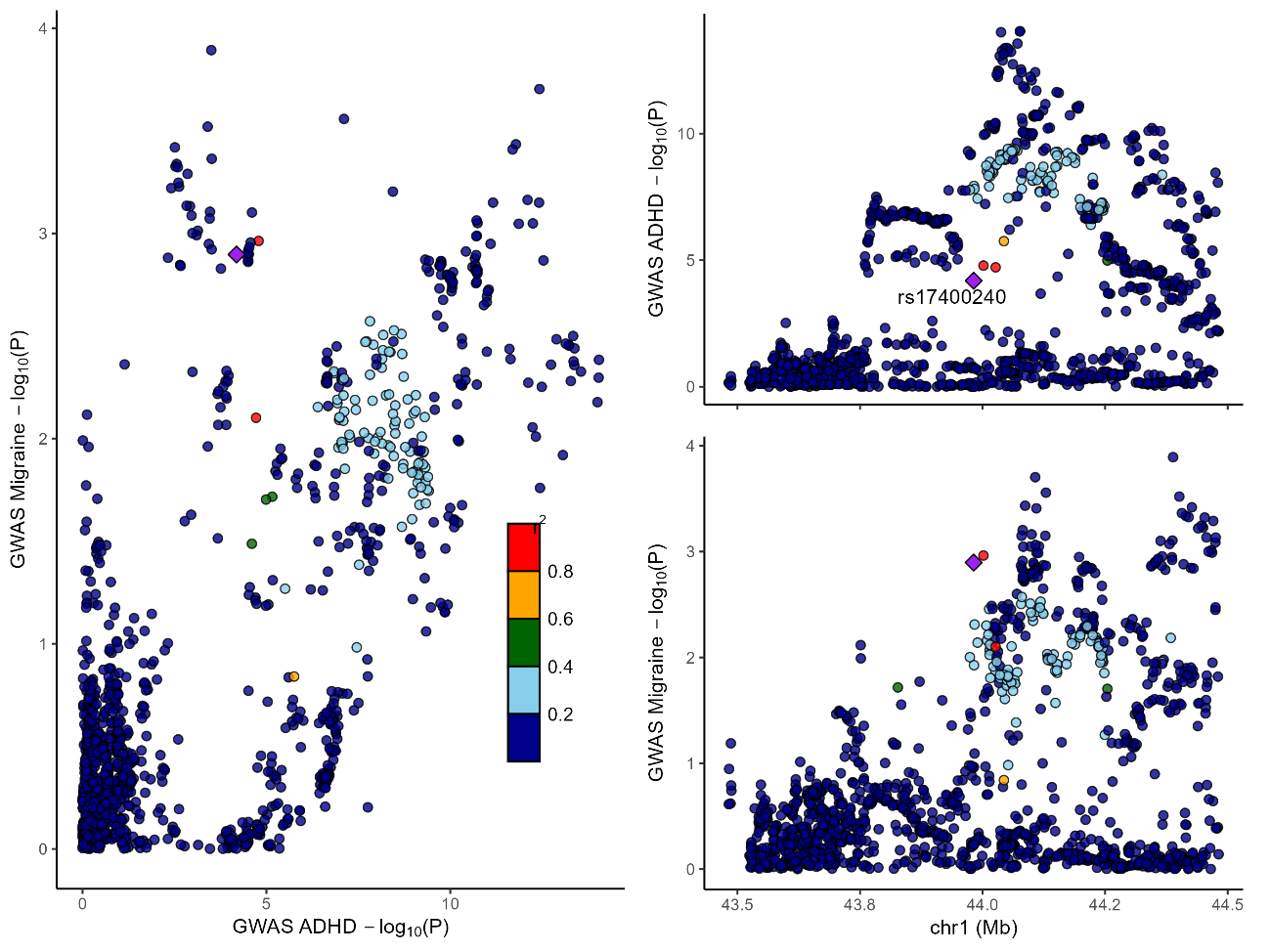

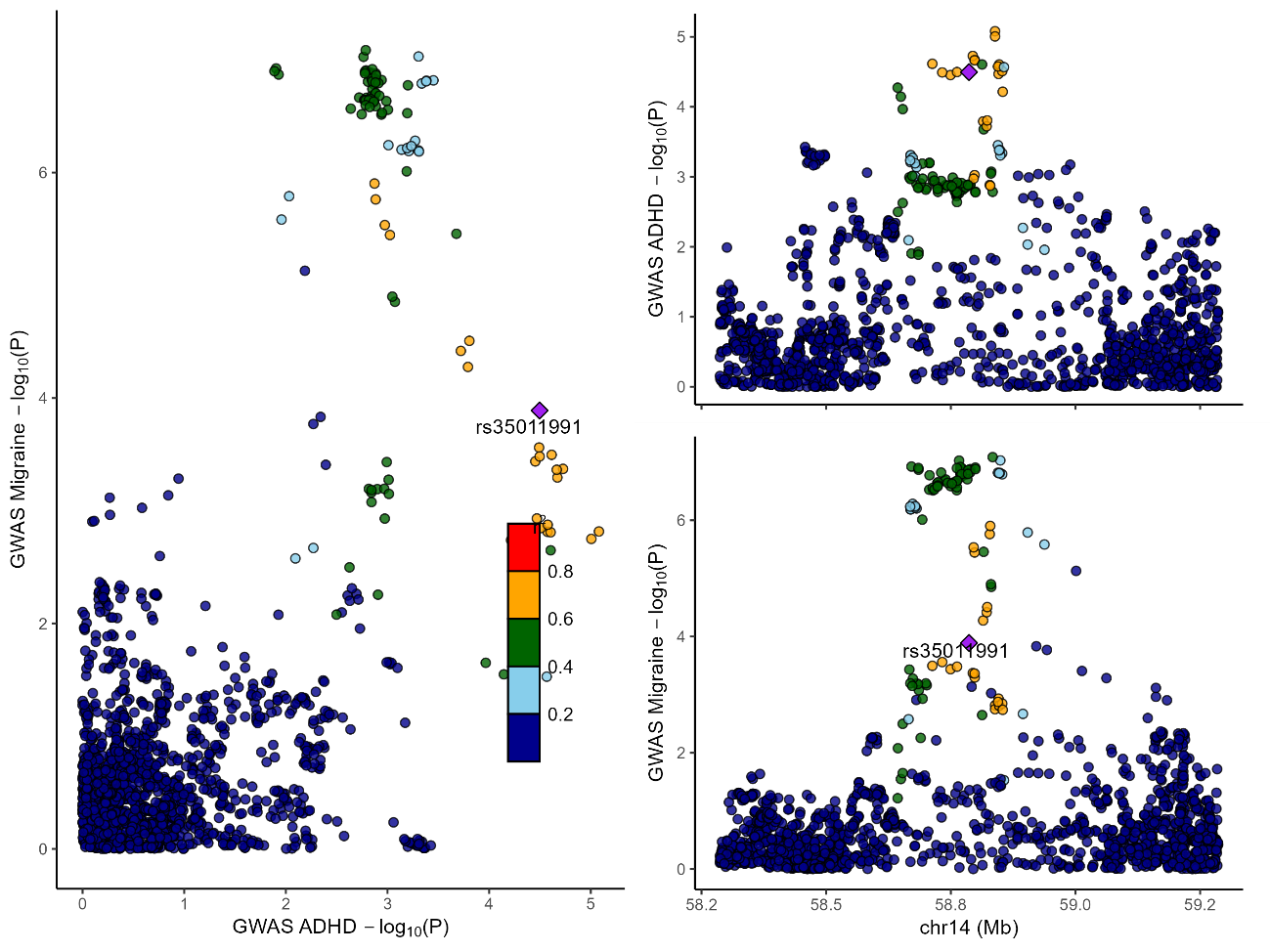

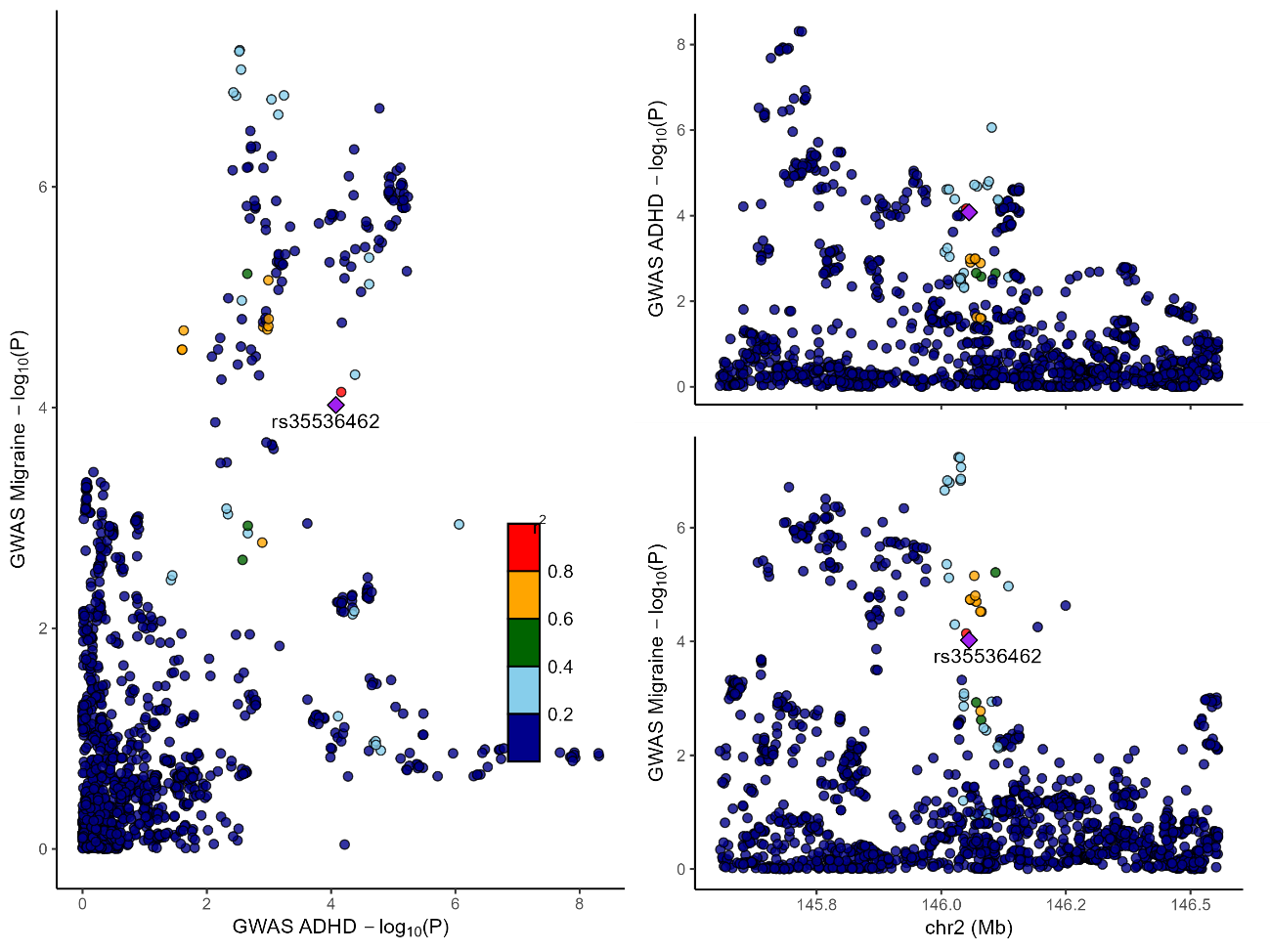

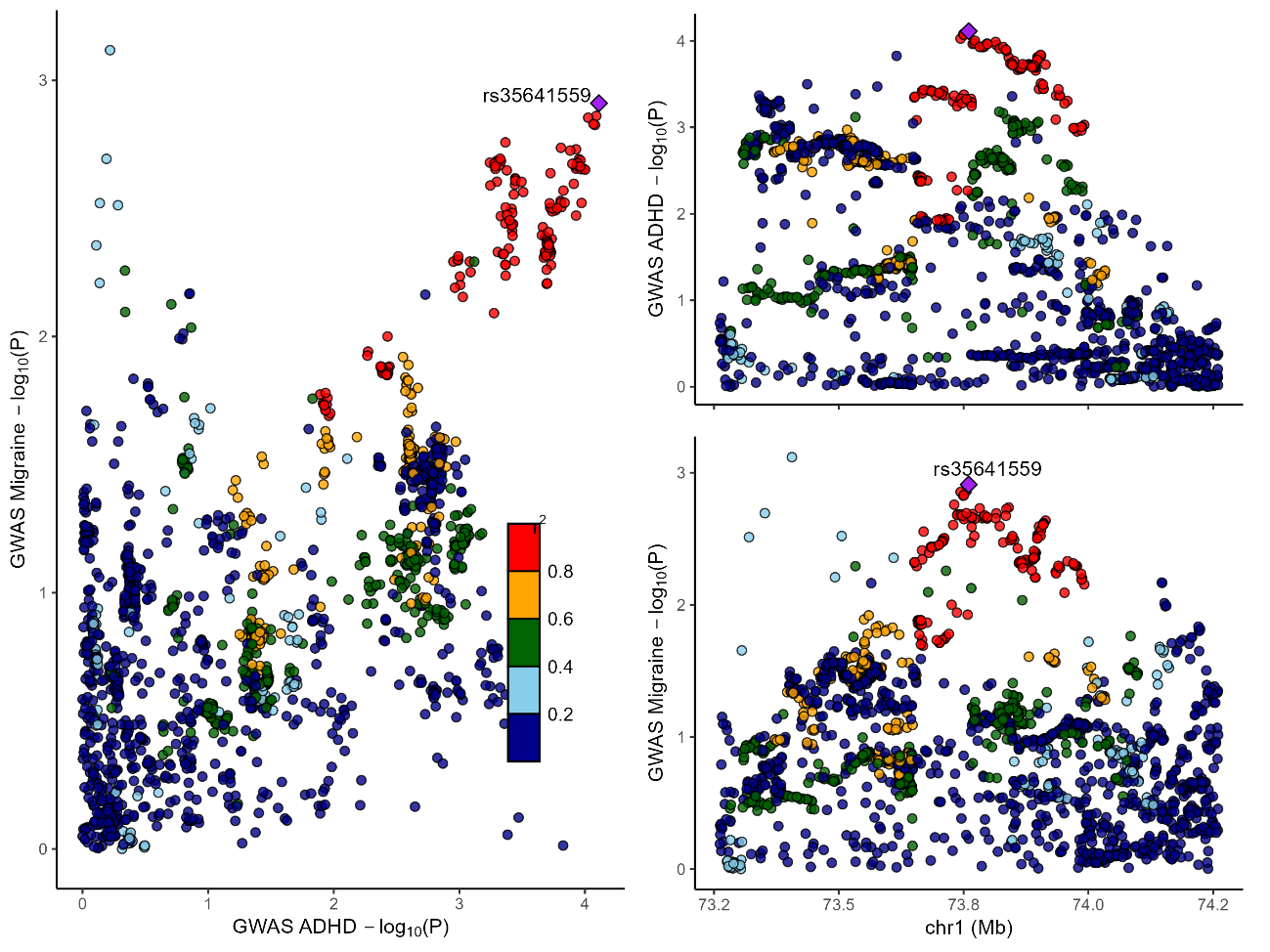

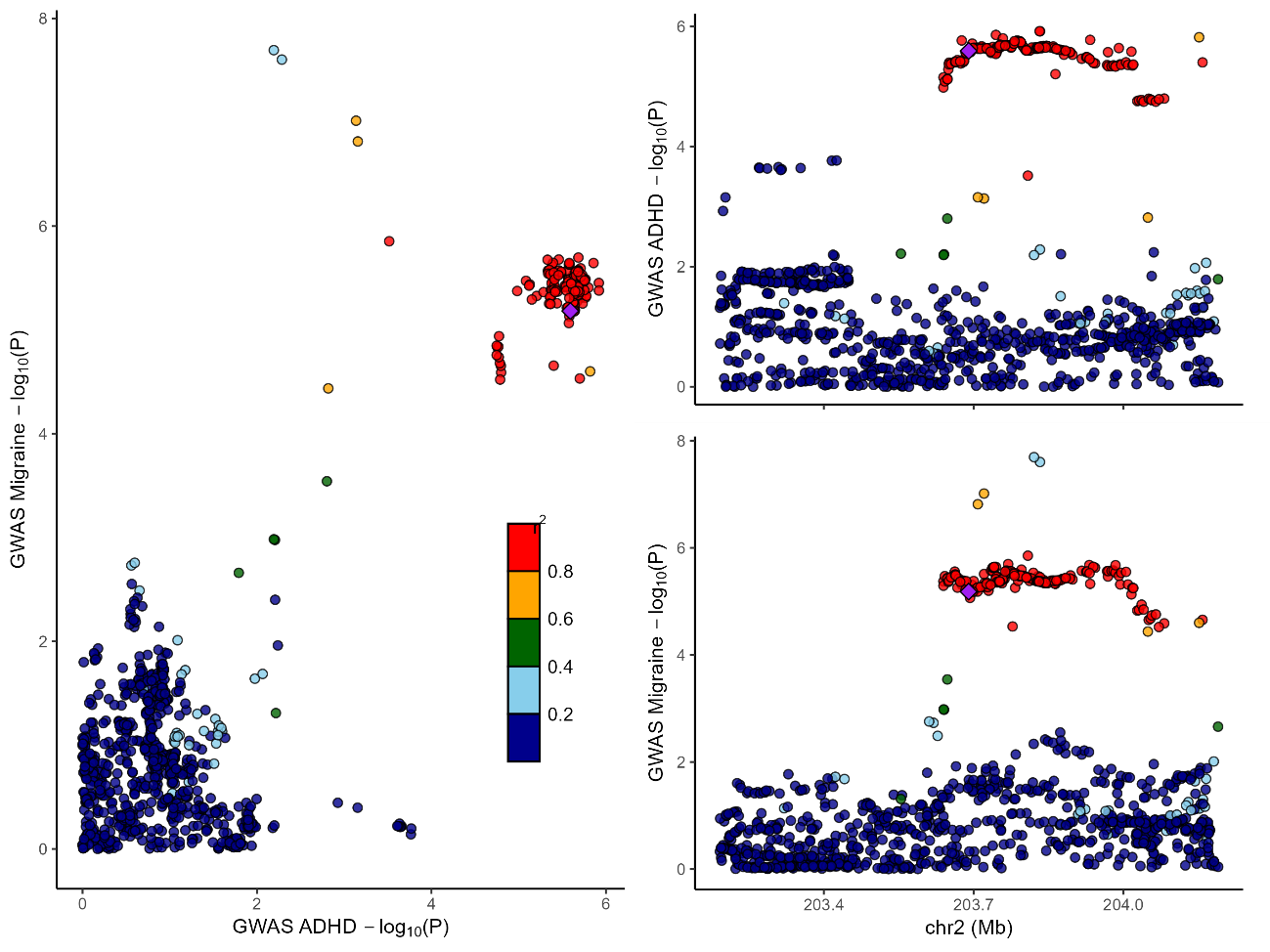

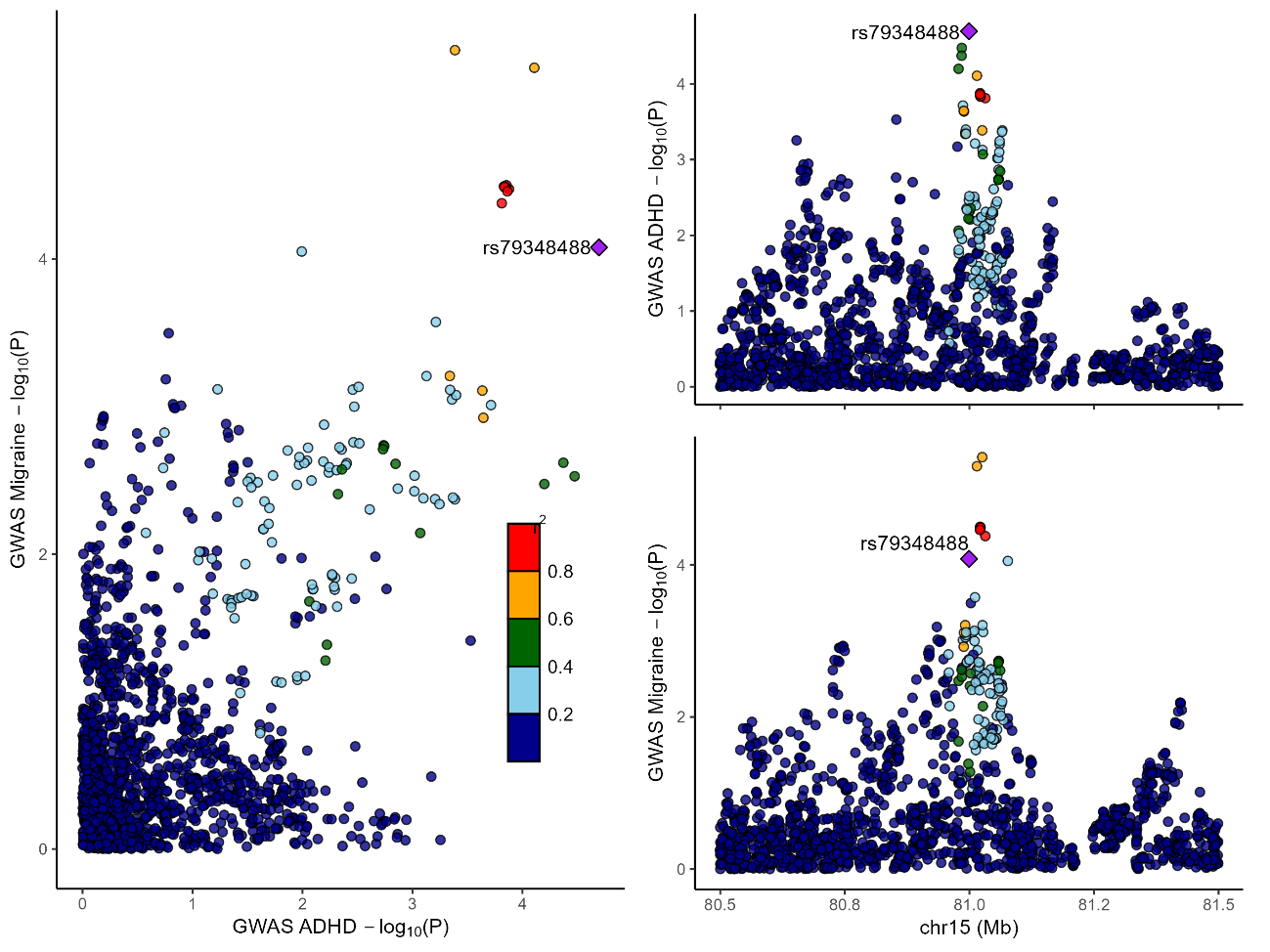

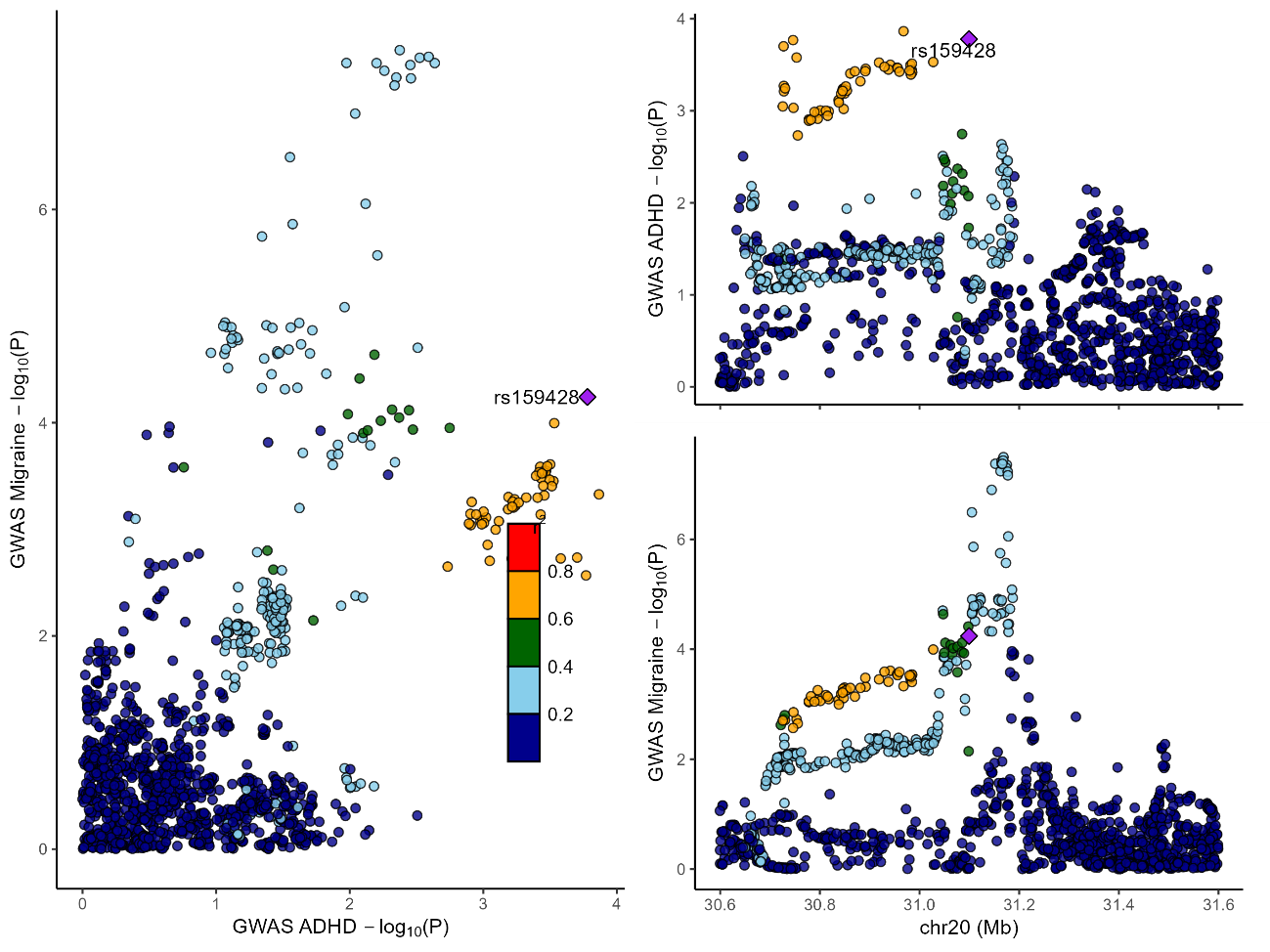

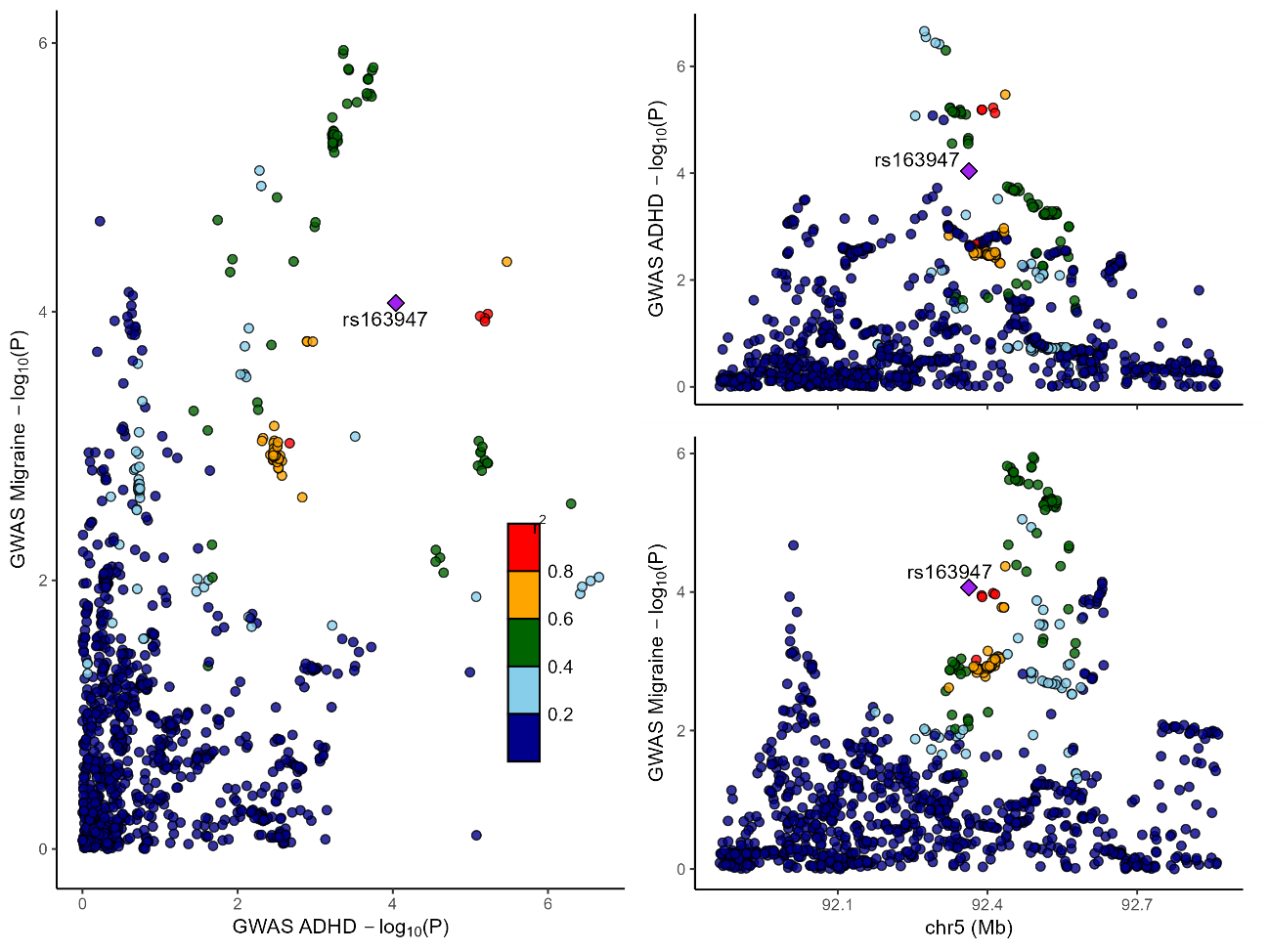

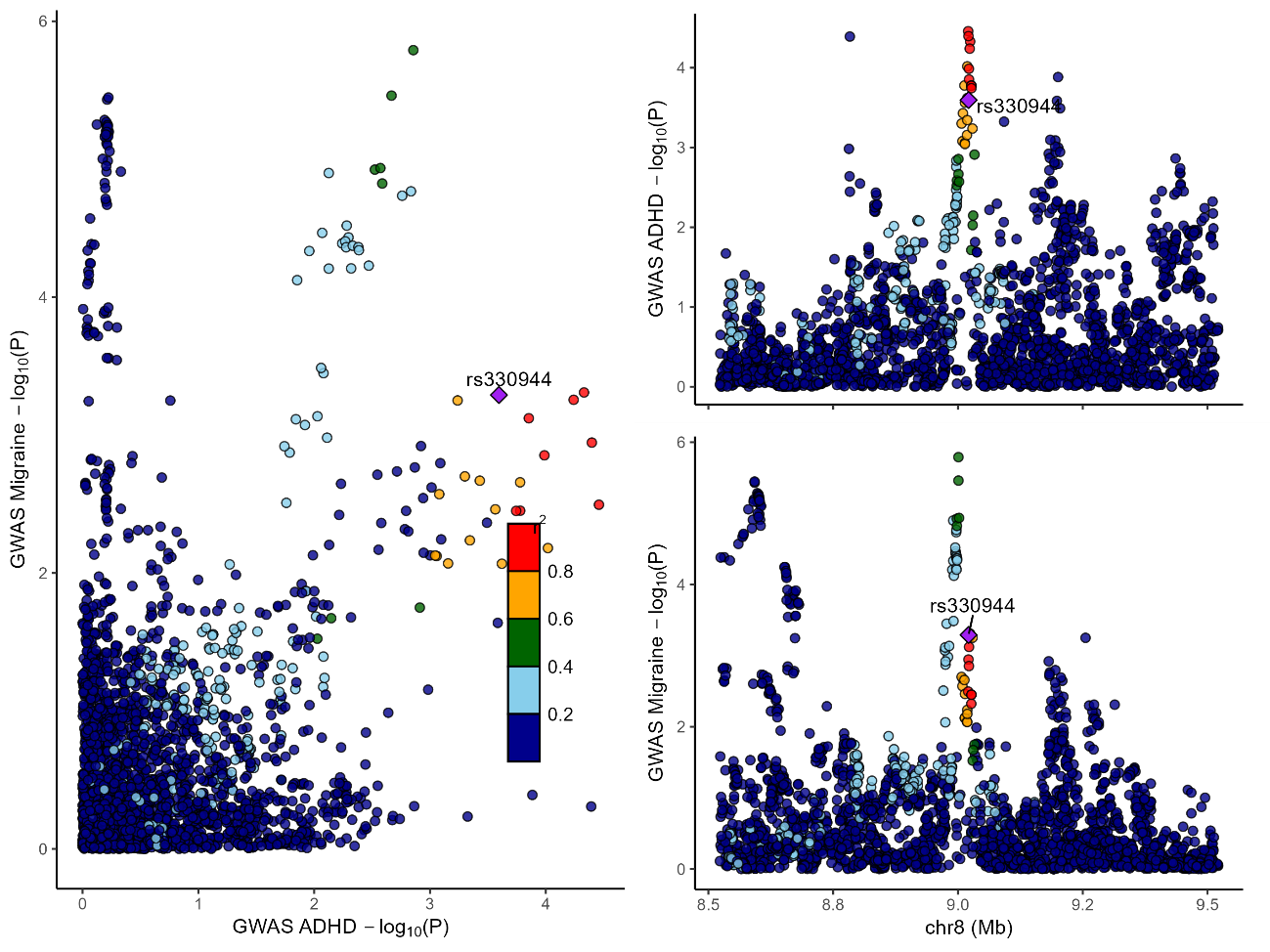

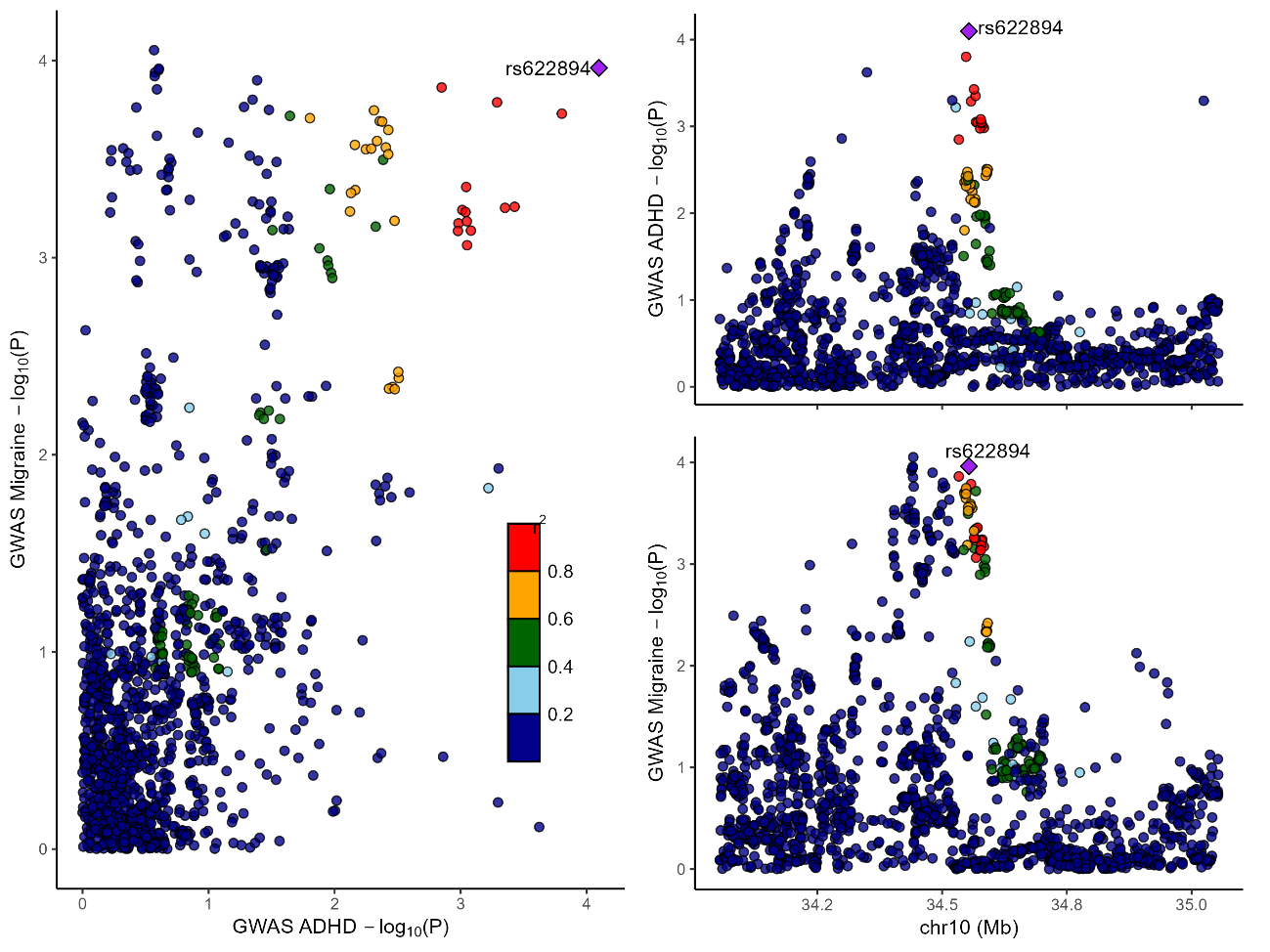

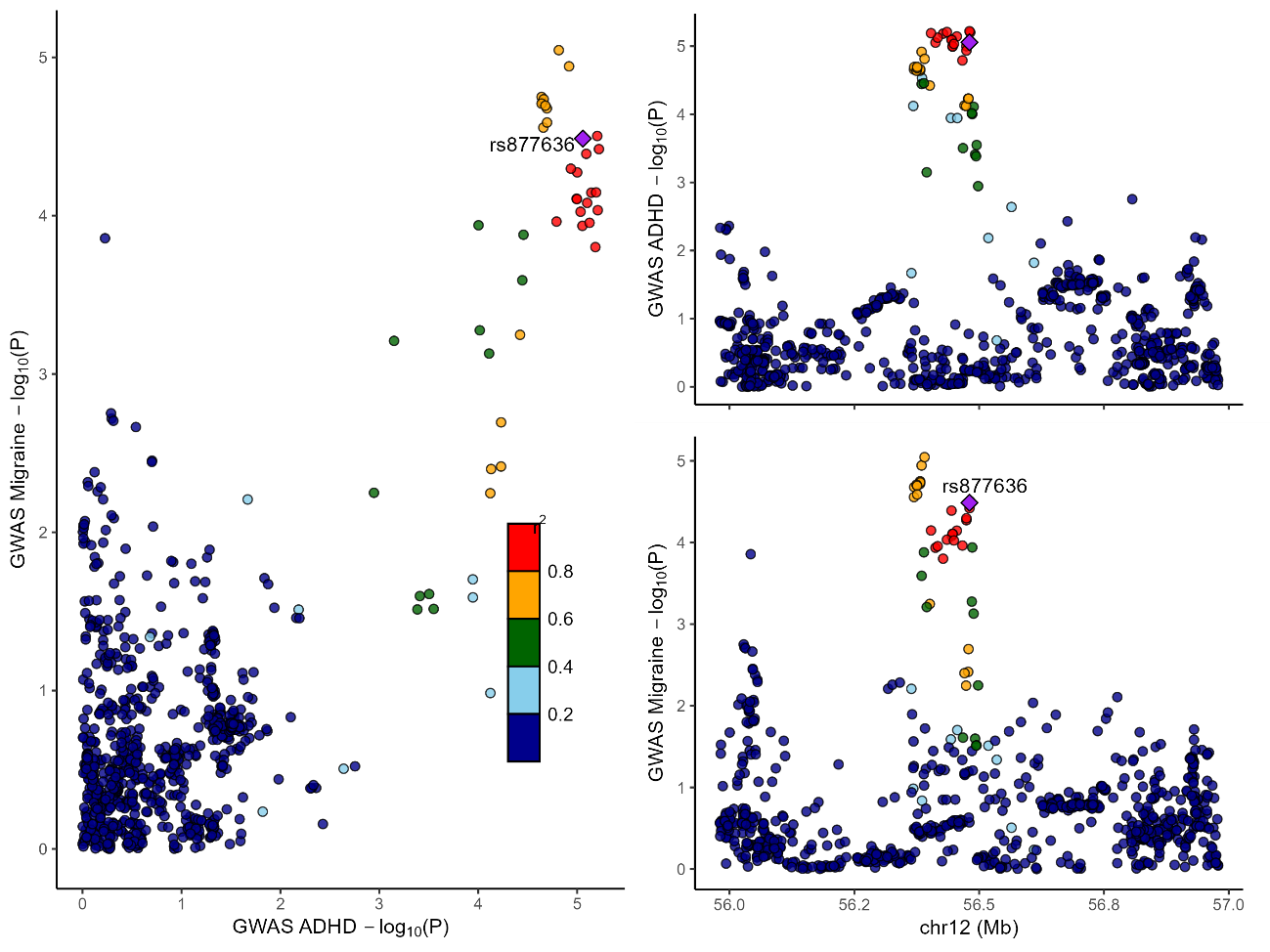

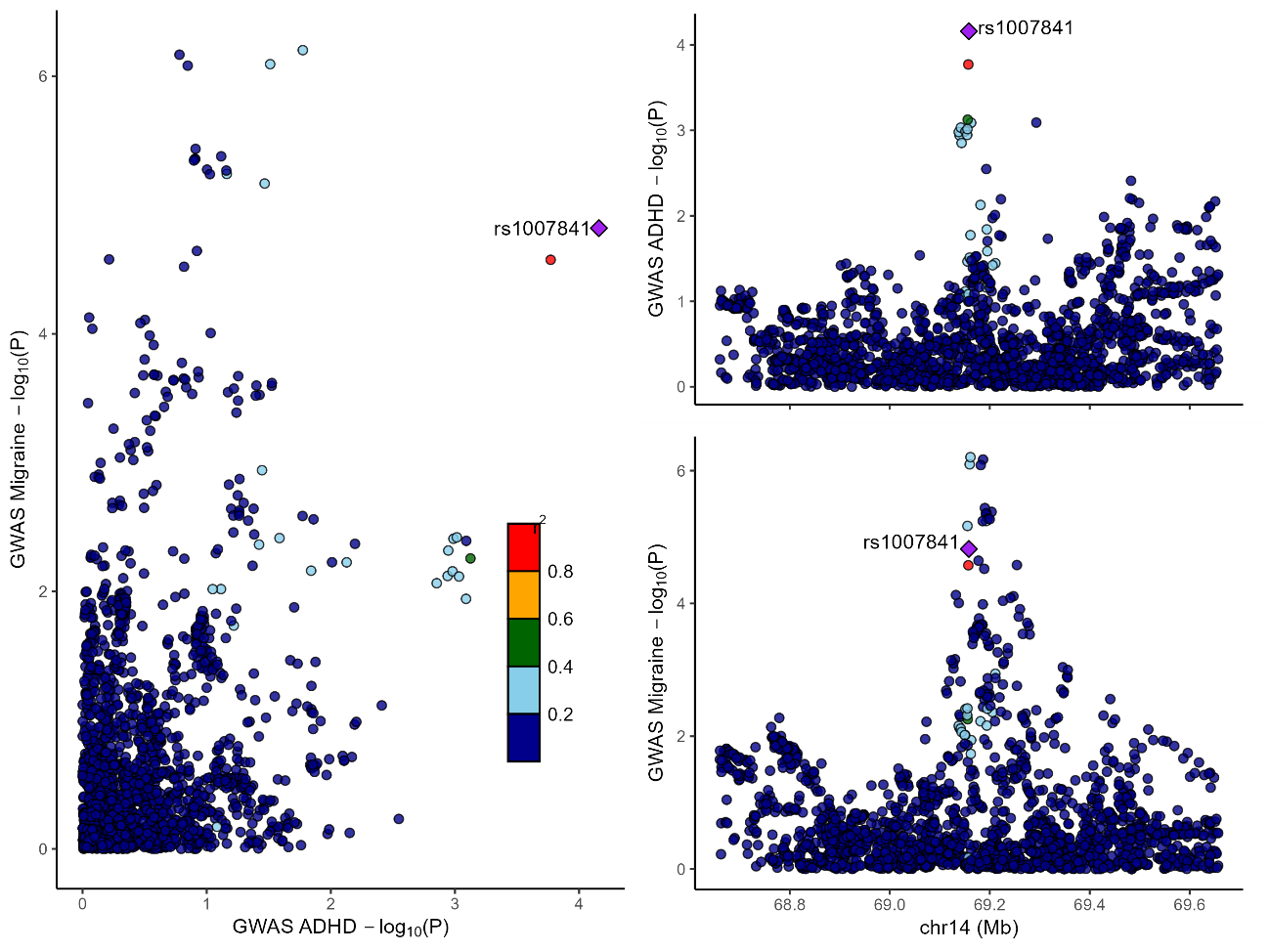

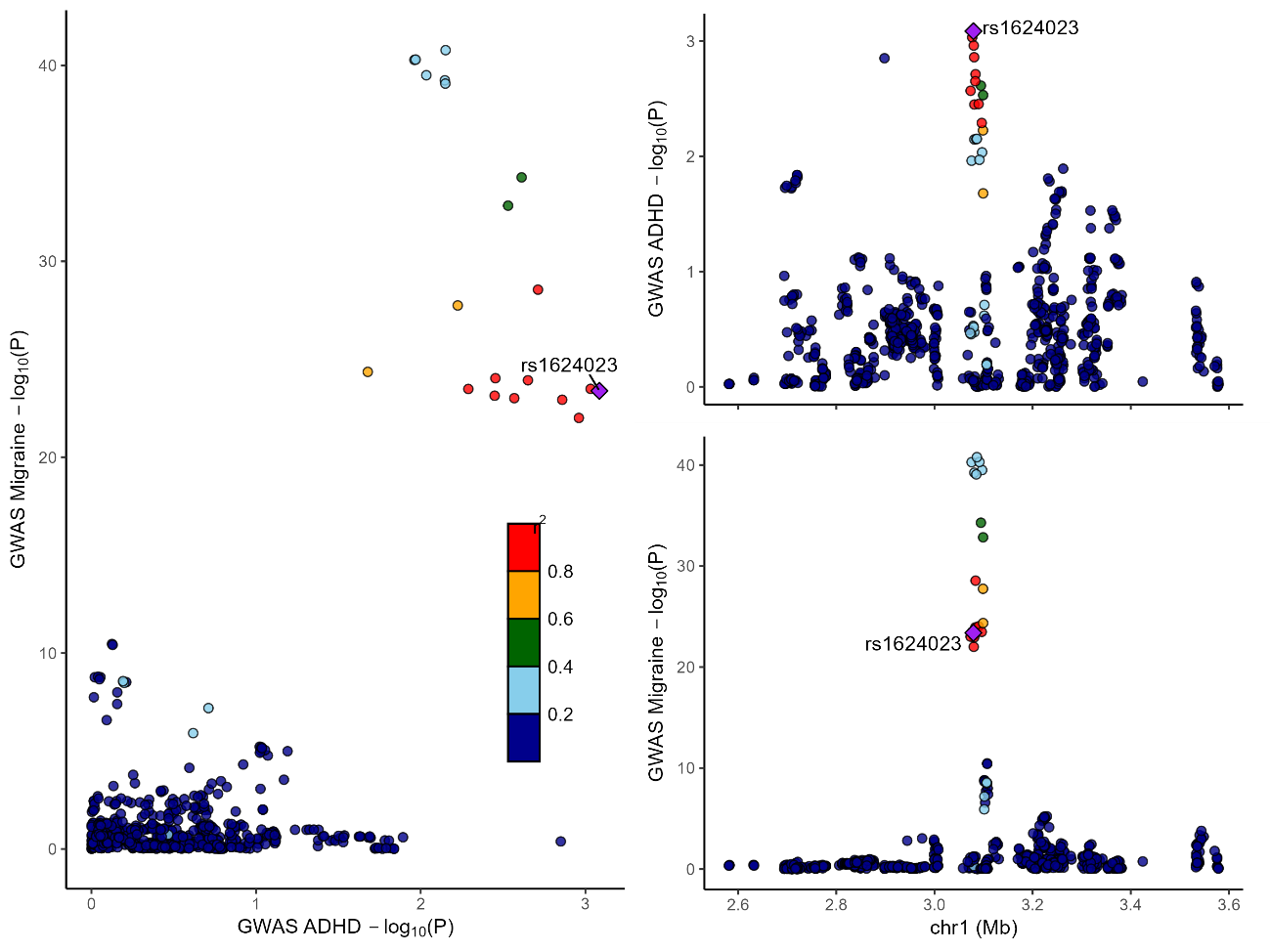

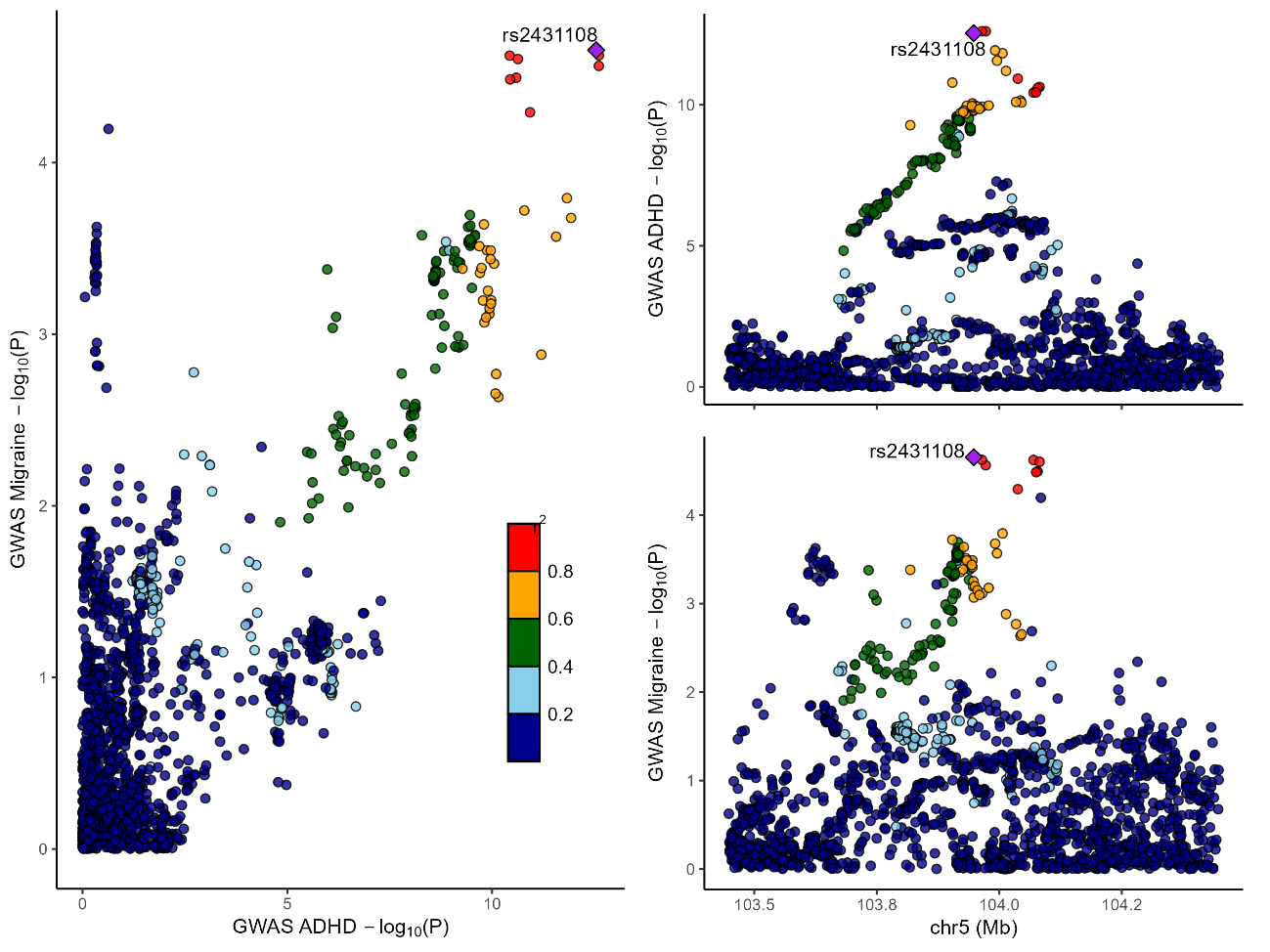

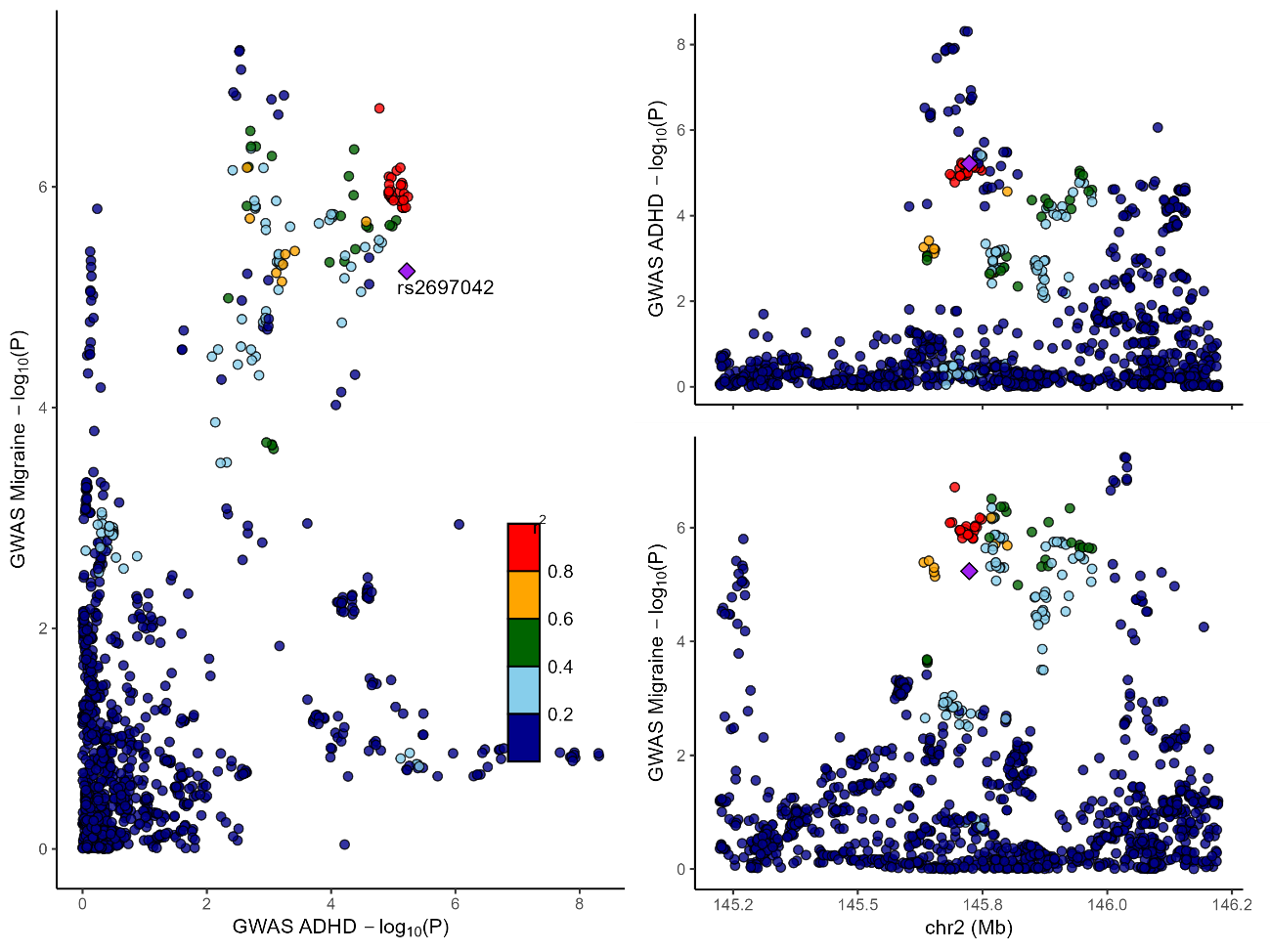

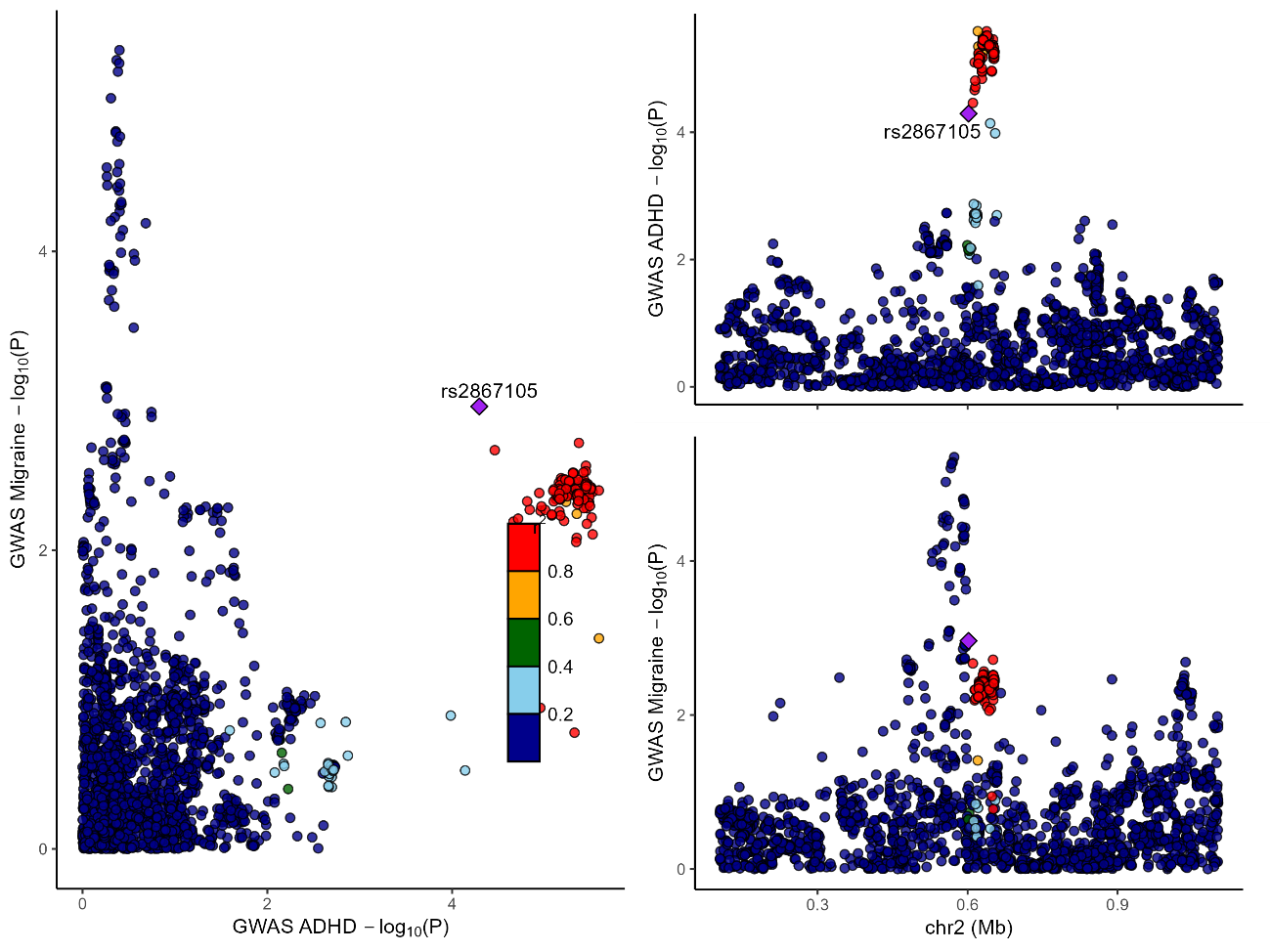

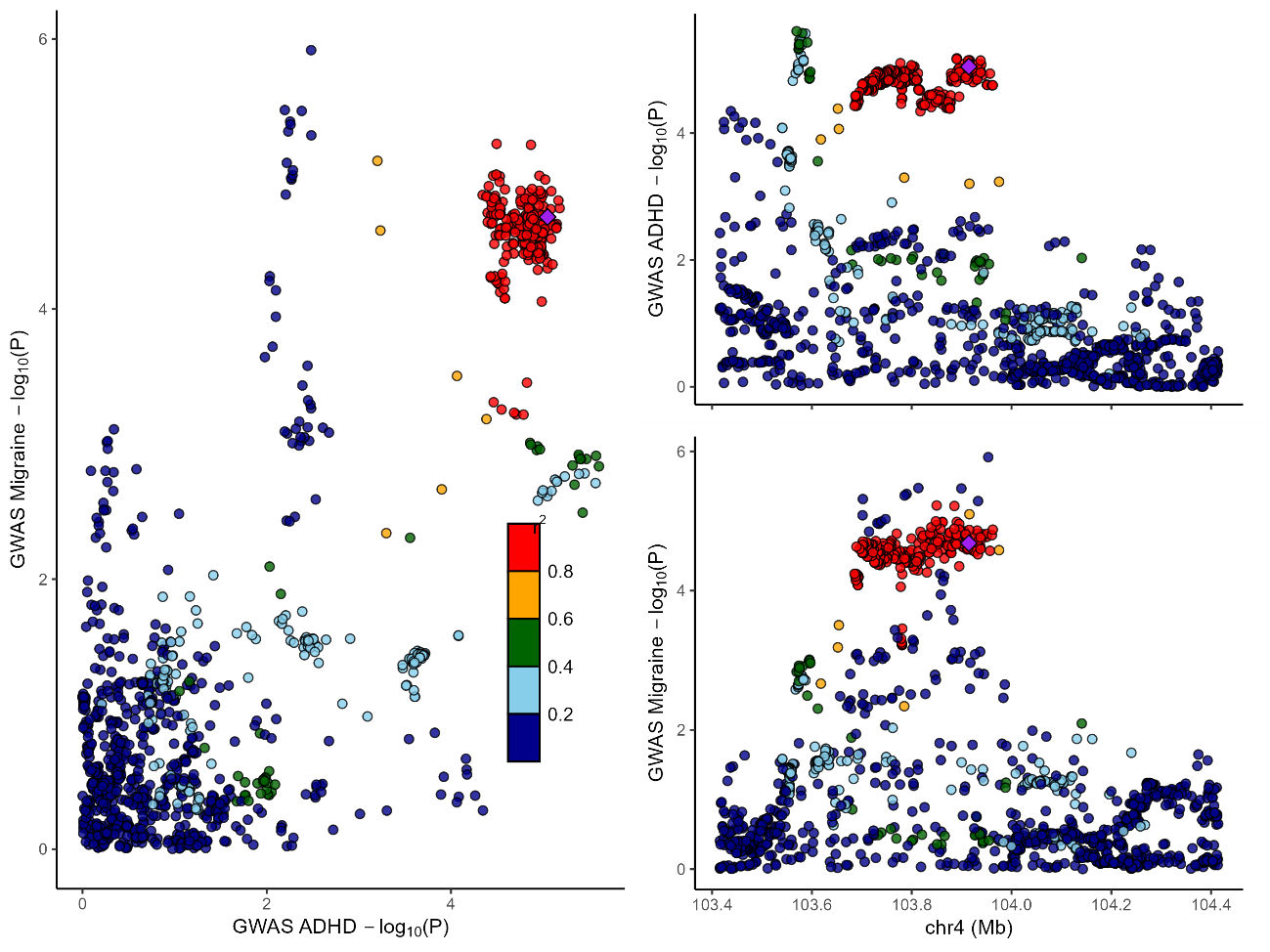

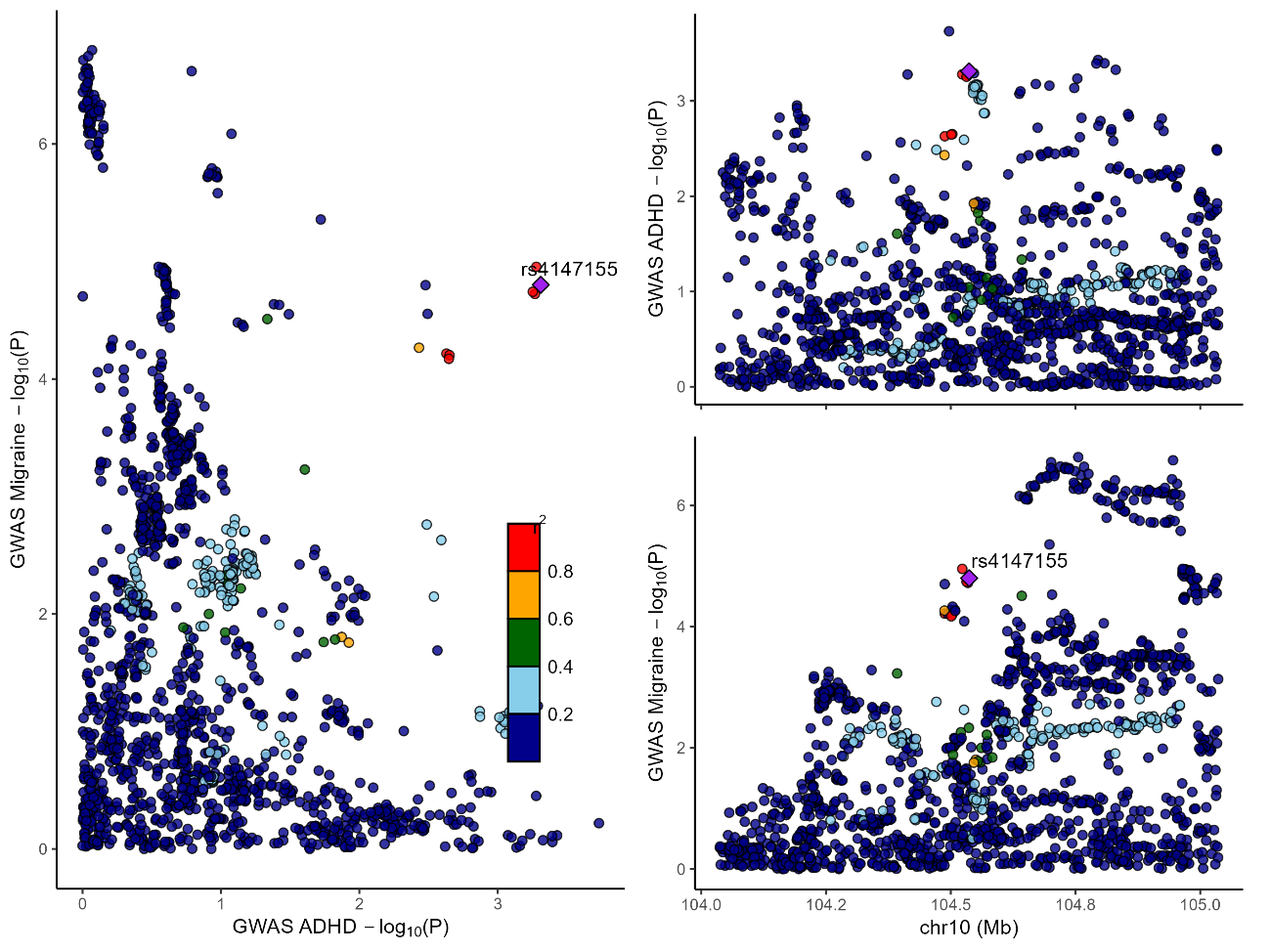

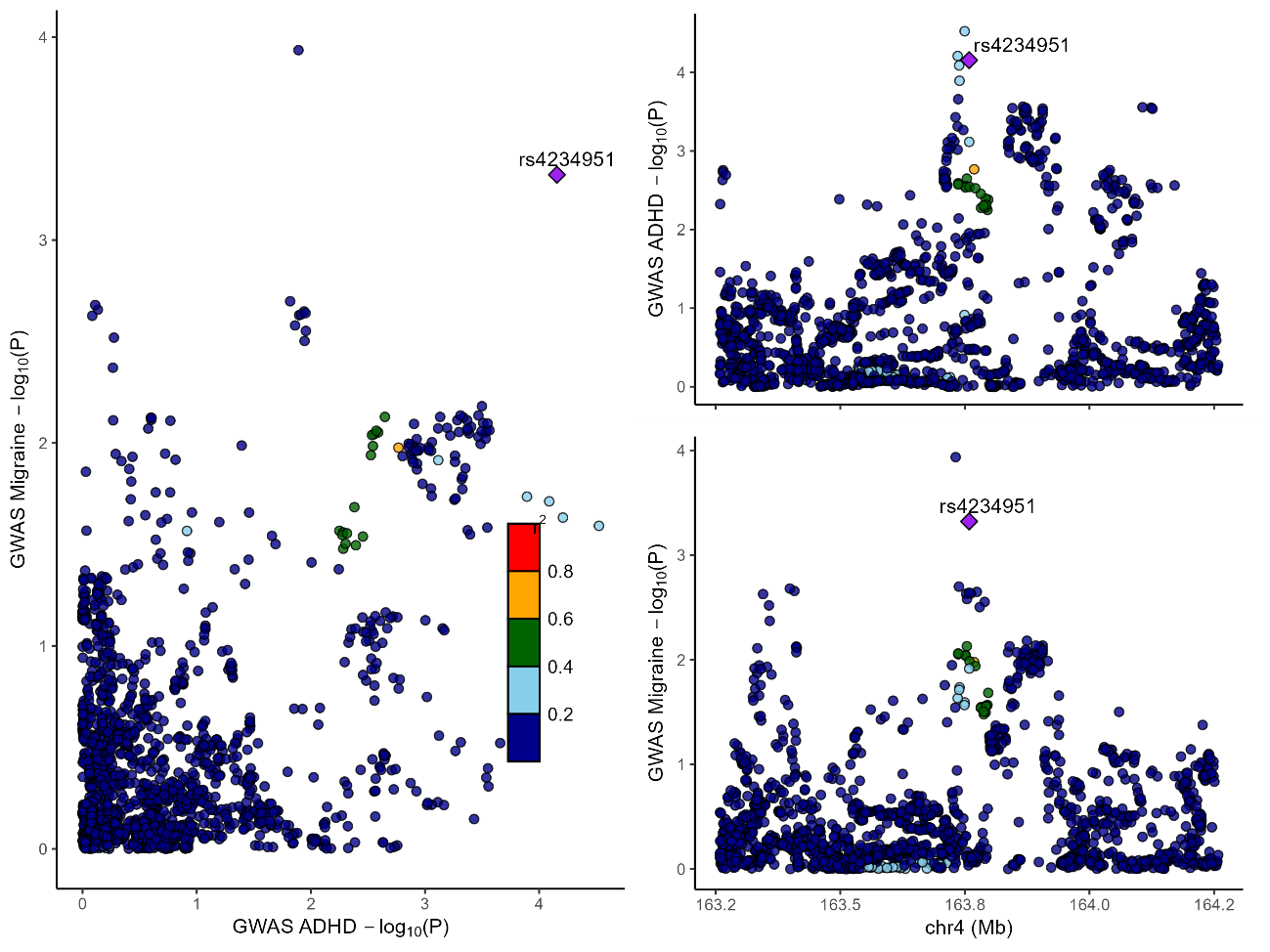

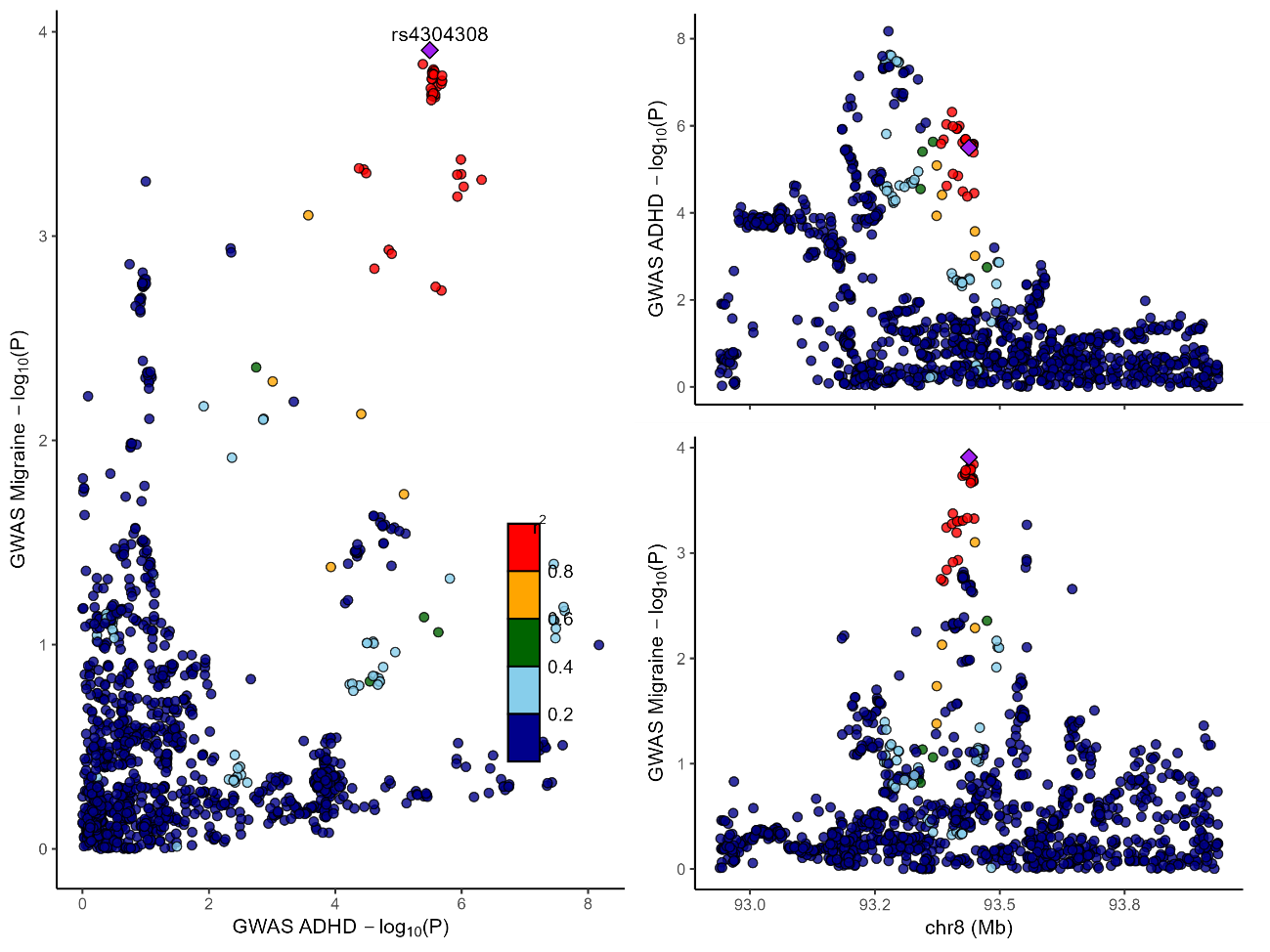

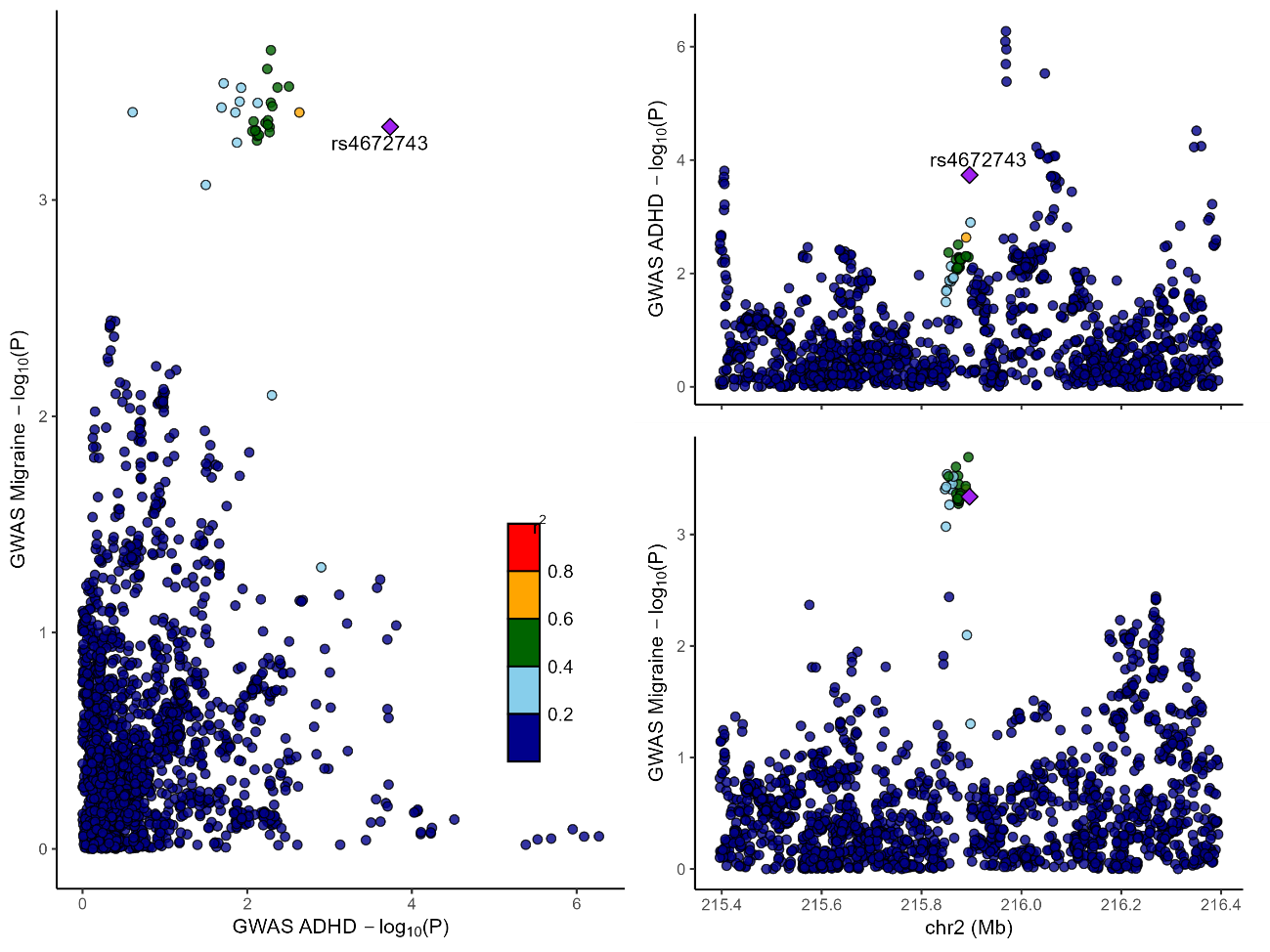

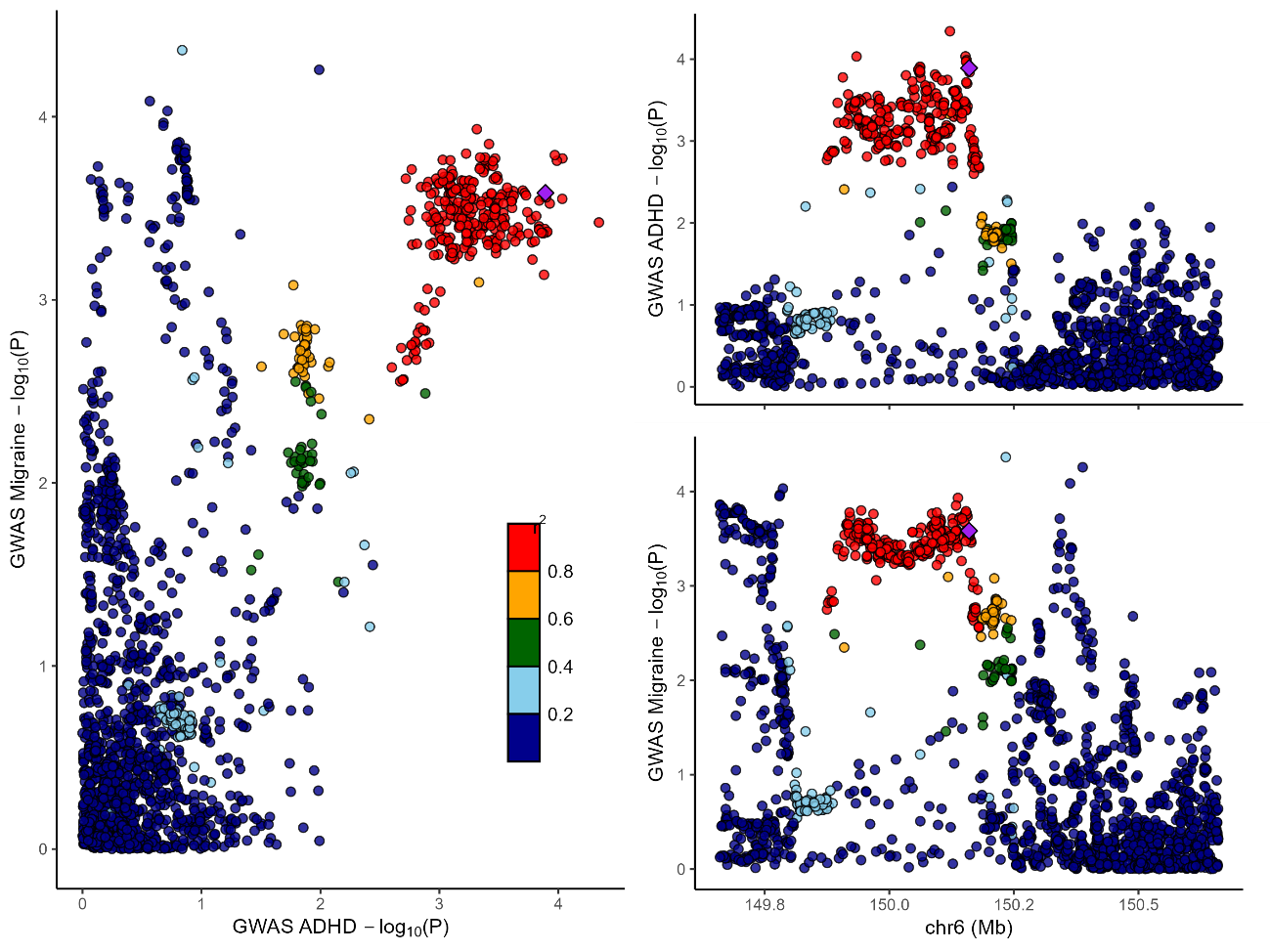

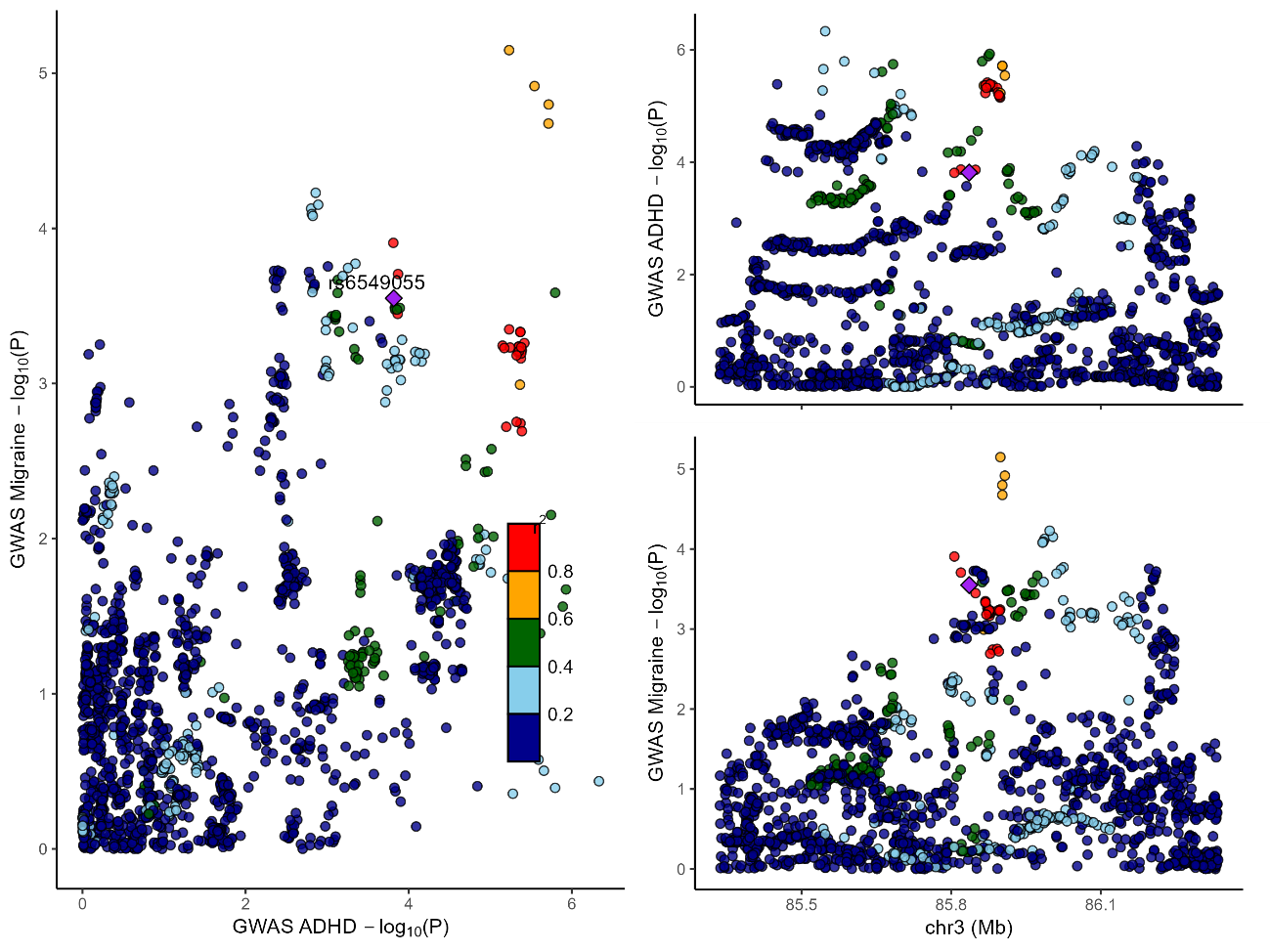

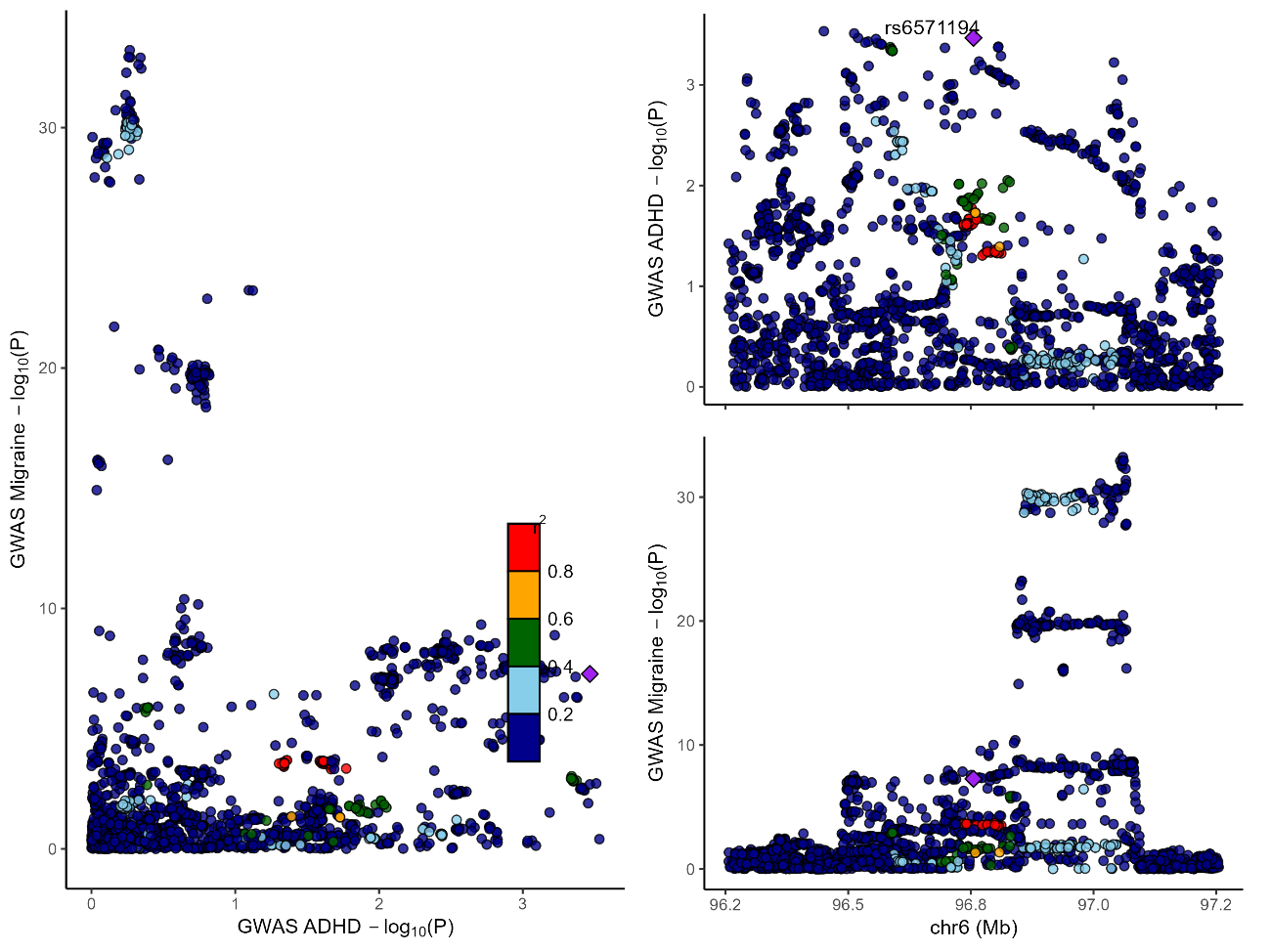

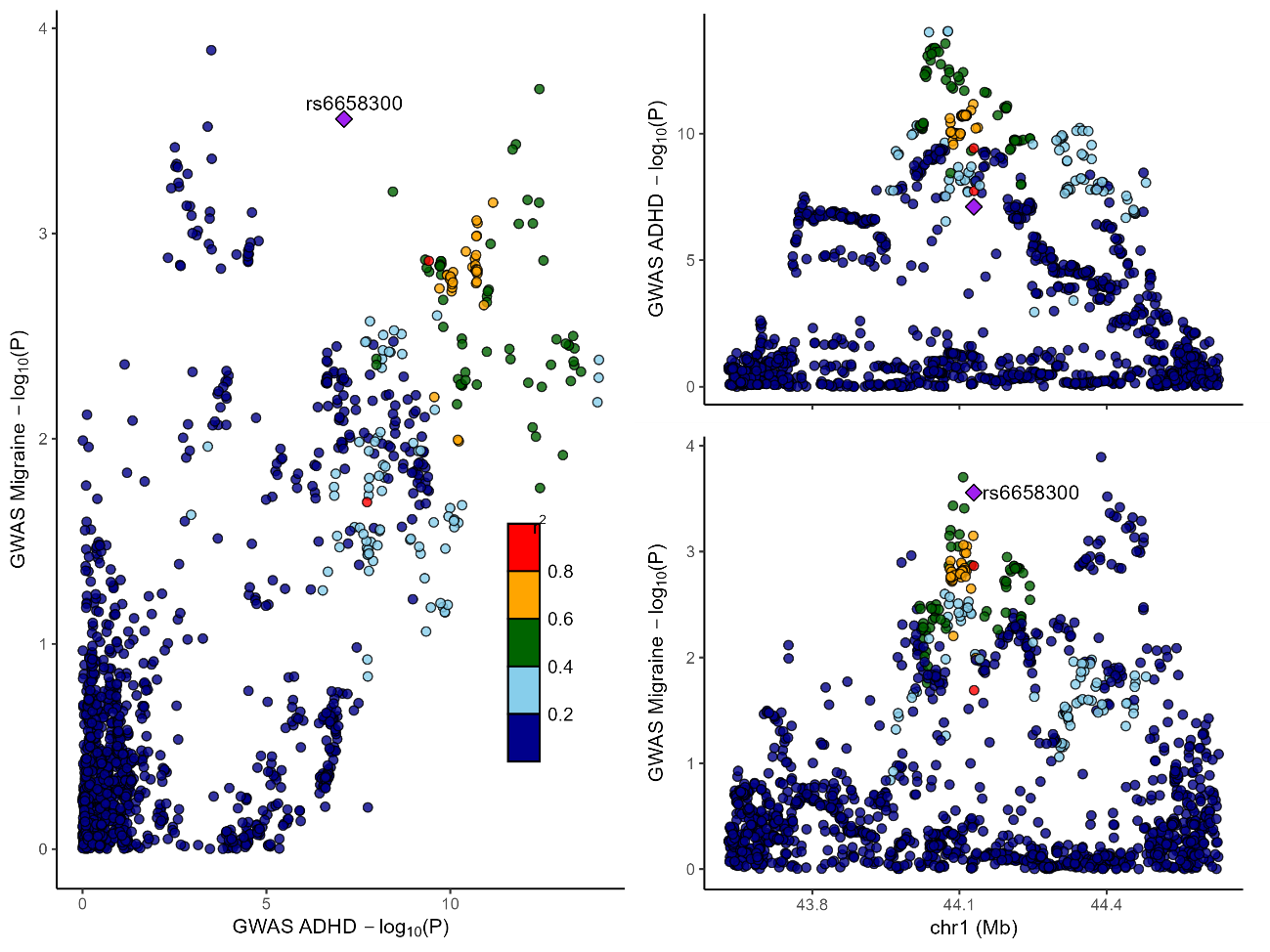

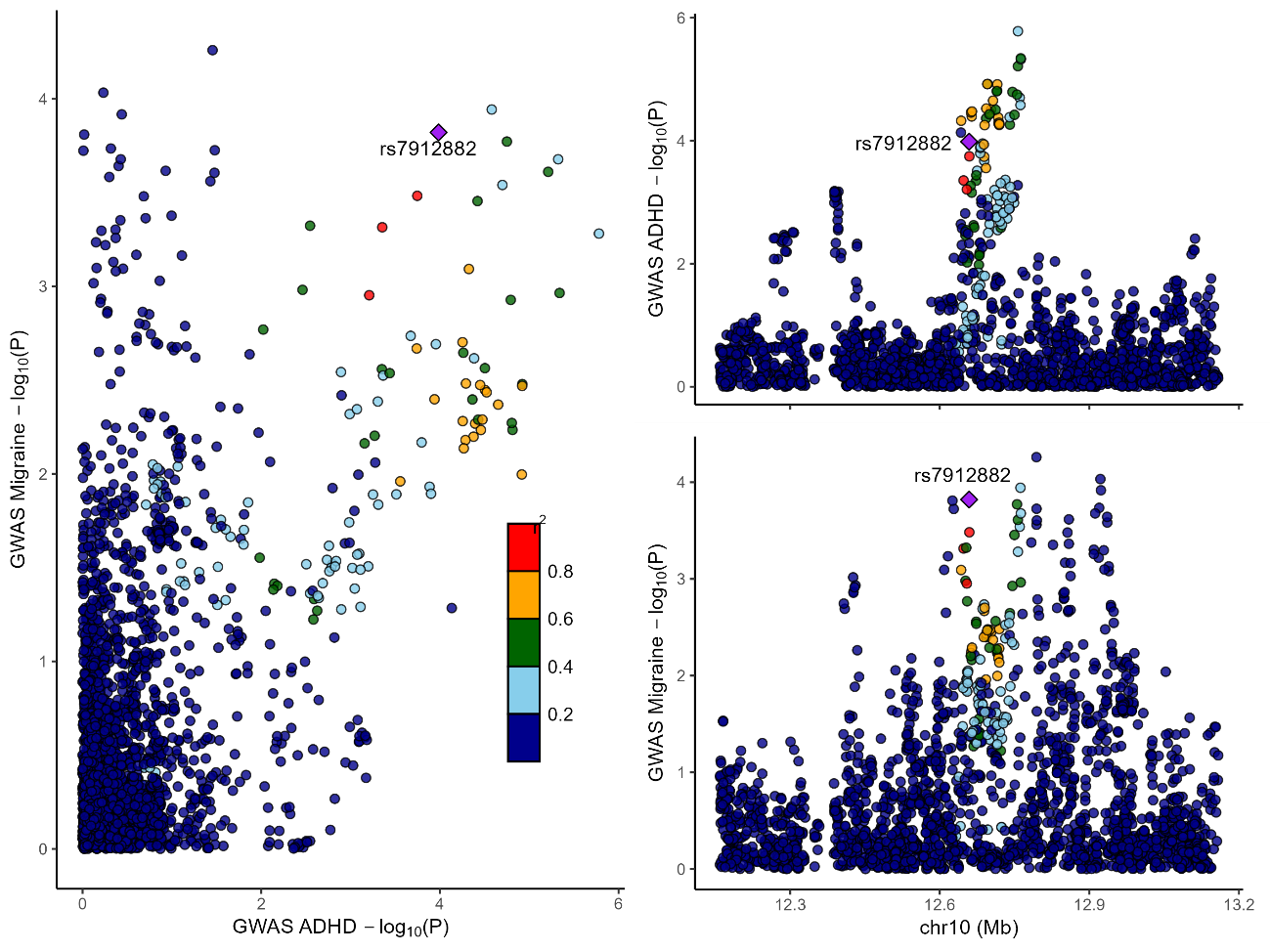

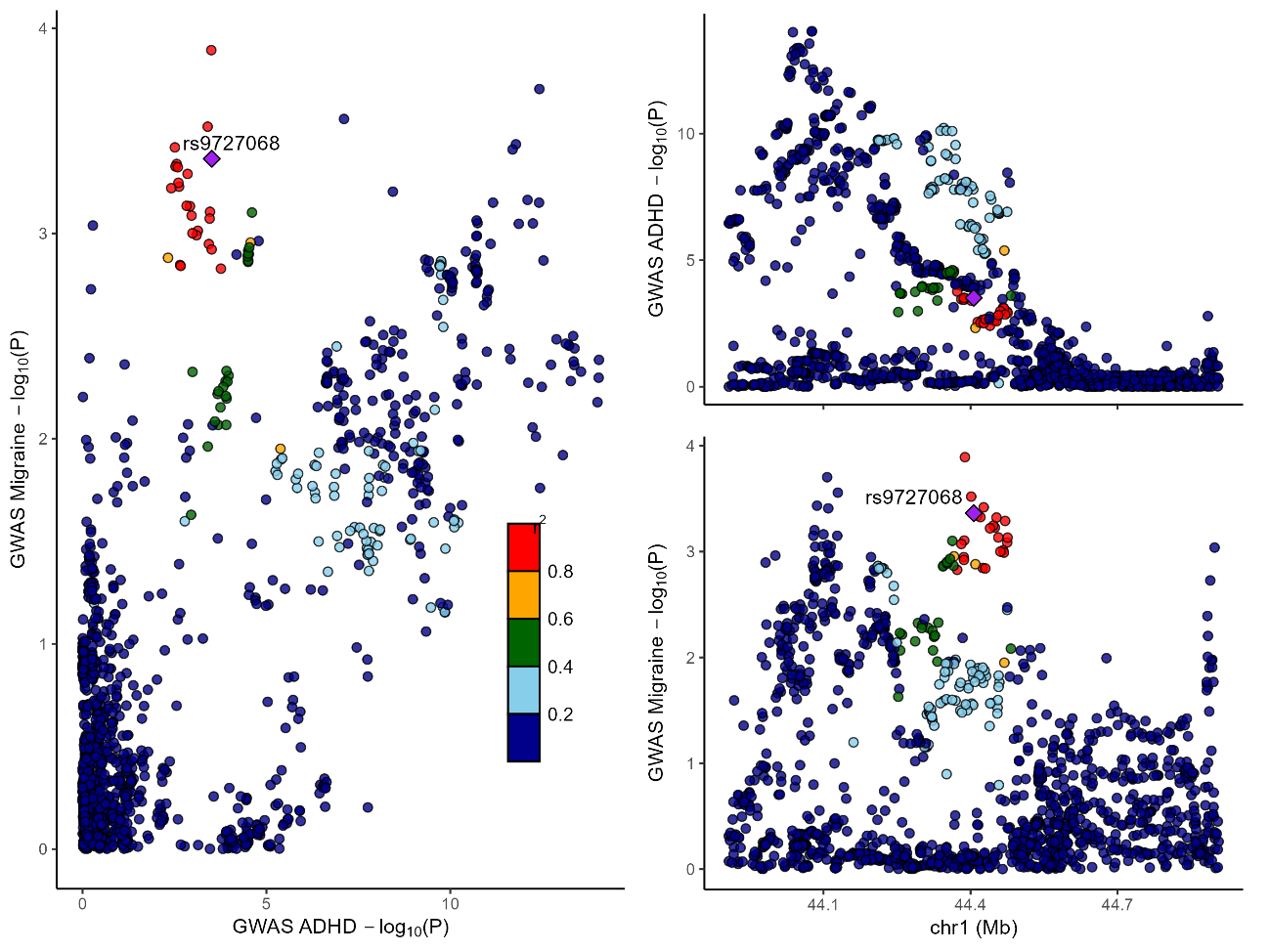

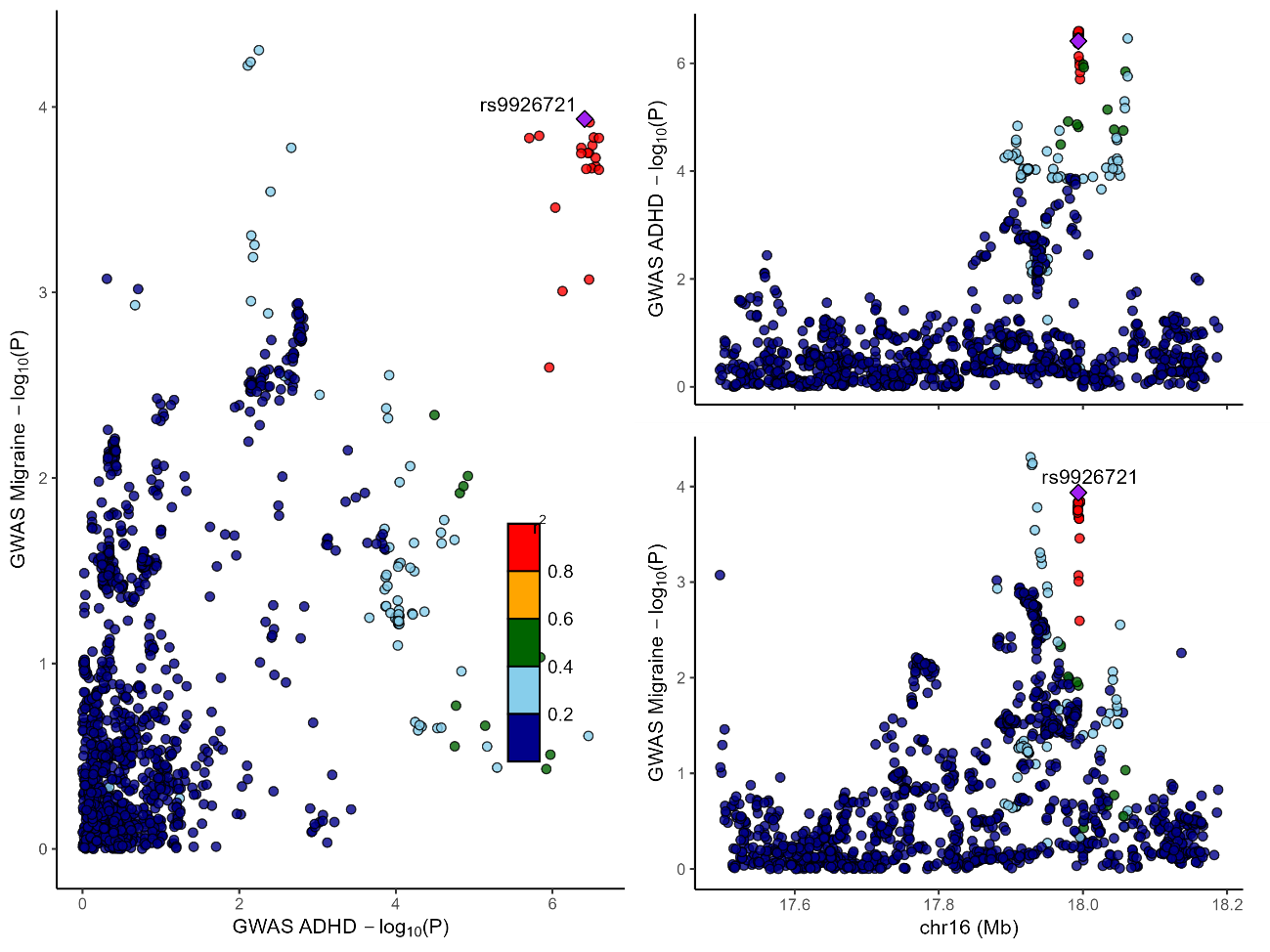

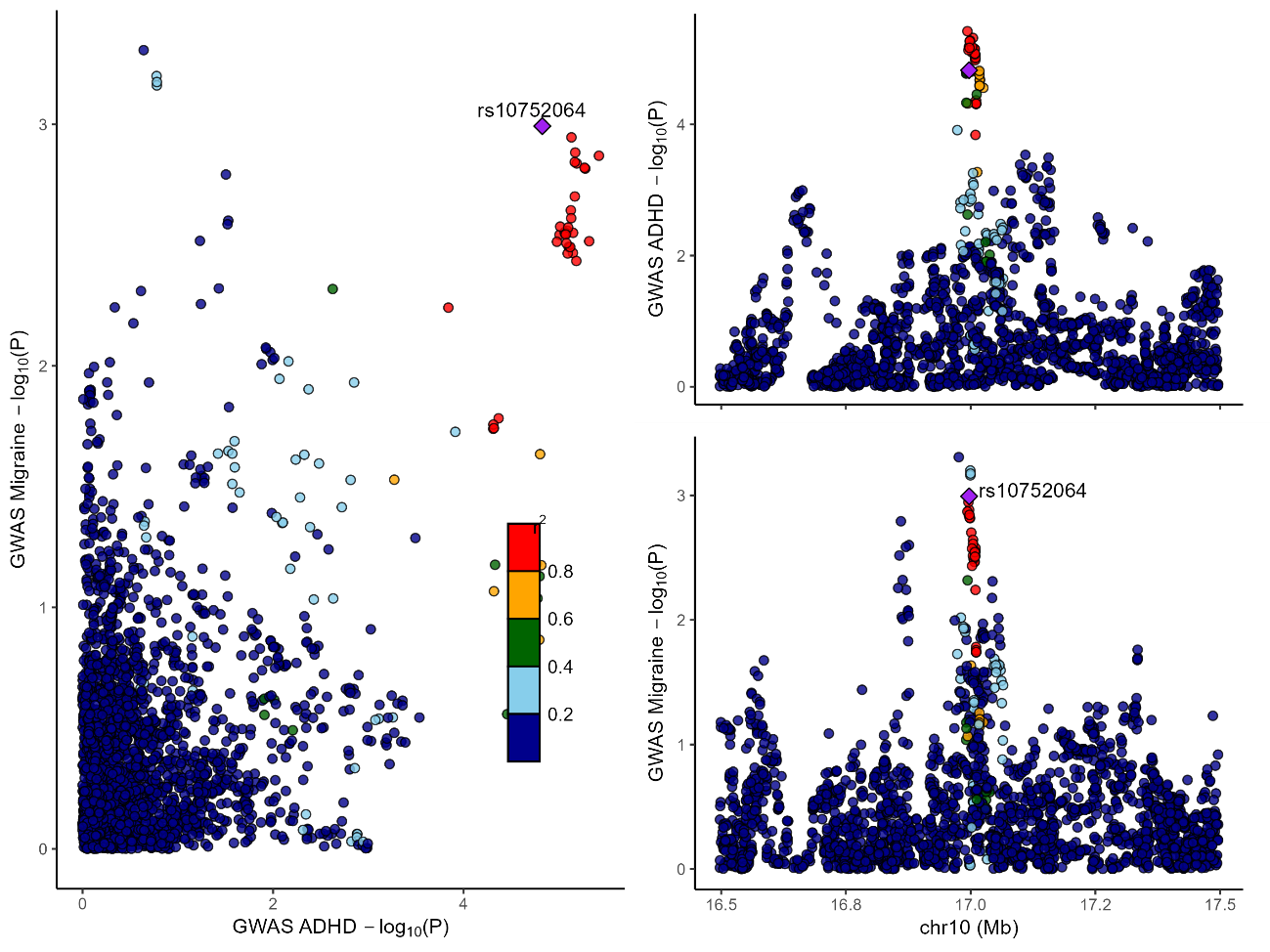

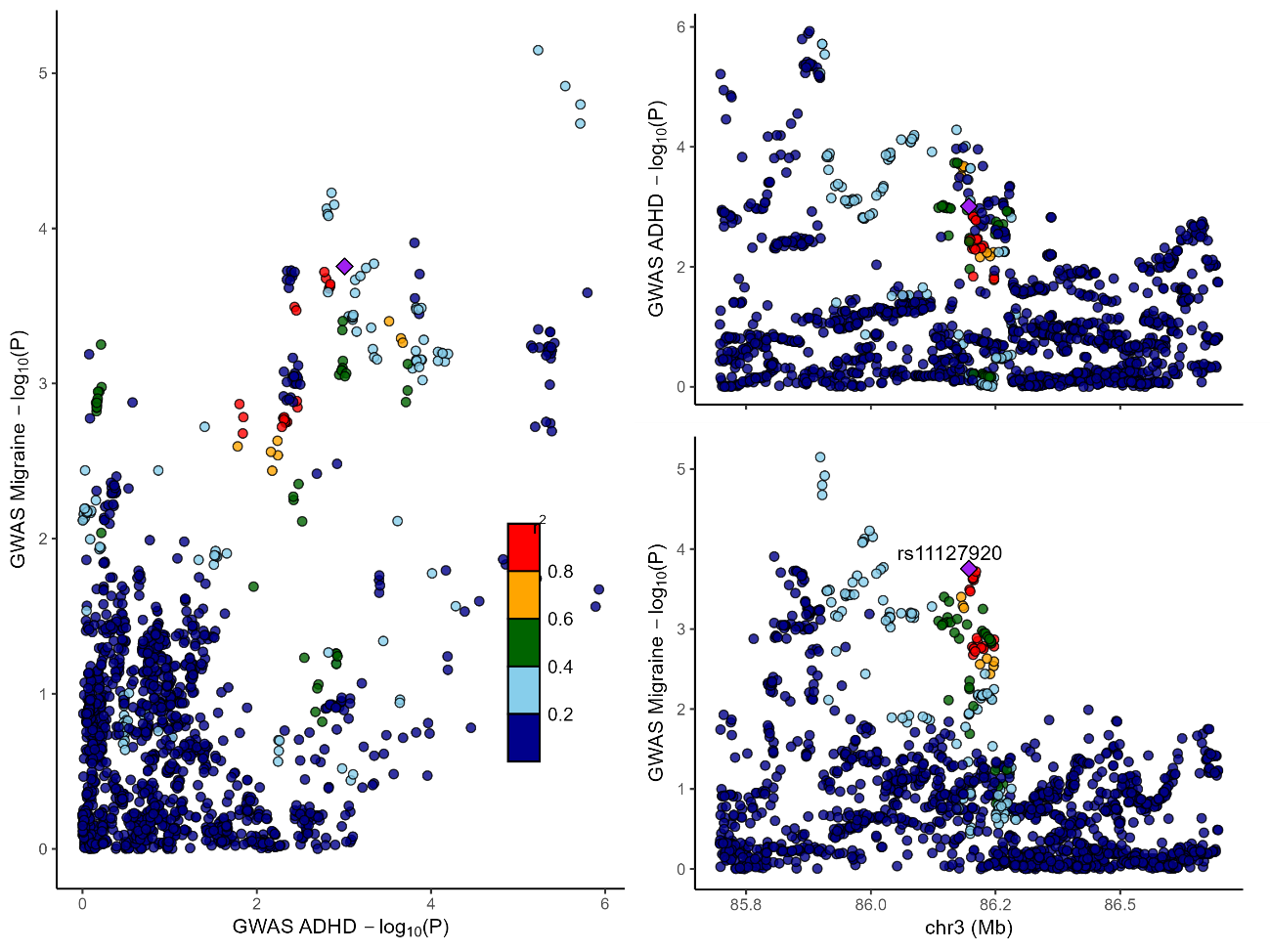

rs72932777

rs72932777

rs72932777

rs3974485

rs3974485

rs4870050

rs4870050

Supplementary Figure 3. Regional plots for each lead SNP from the cross-trait analysis of ADHD and migraine. For each variant, the figure includes: (i)Scatter plot (left) relating -log_10_(P) of ADHD (x-axis) and migraine (y-axis) (ii) Region plots (right) displaying regional SNPs and its association for each trait, ADHD (upper right) and migraine (lower right). Each dot represents a SNP, and colour scale indicates linkage disequilibrium (LD; r2) with the top variant, highlighted as a purple diamond.

D)

C)

B)

A)

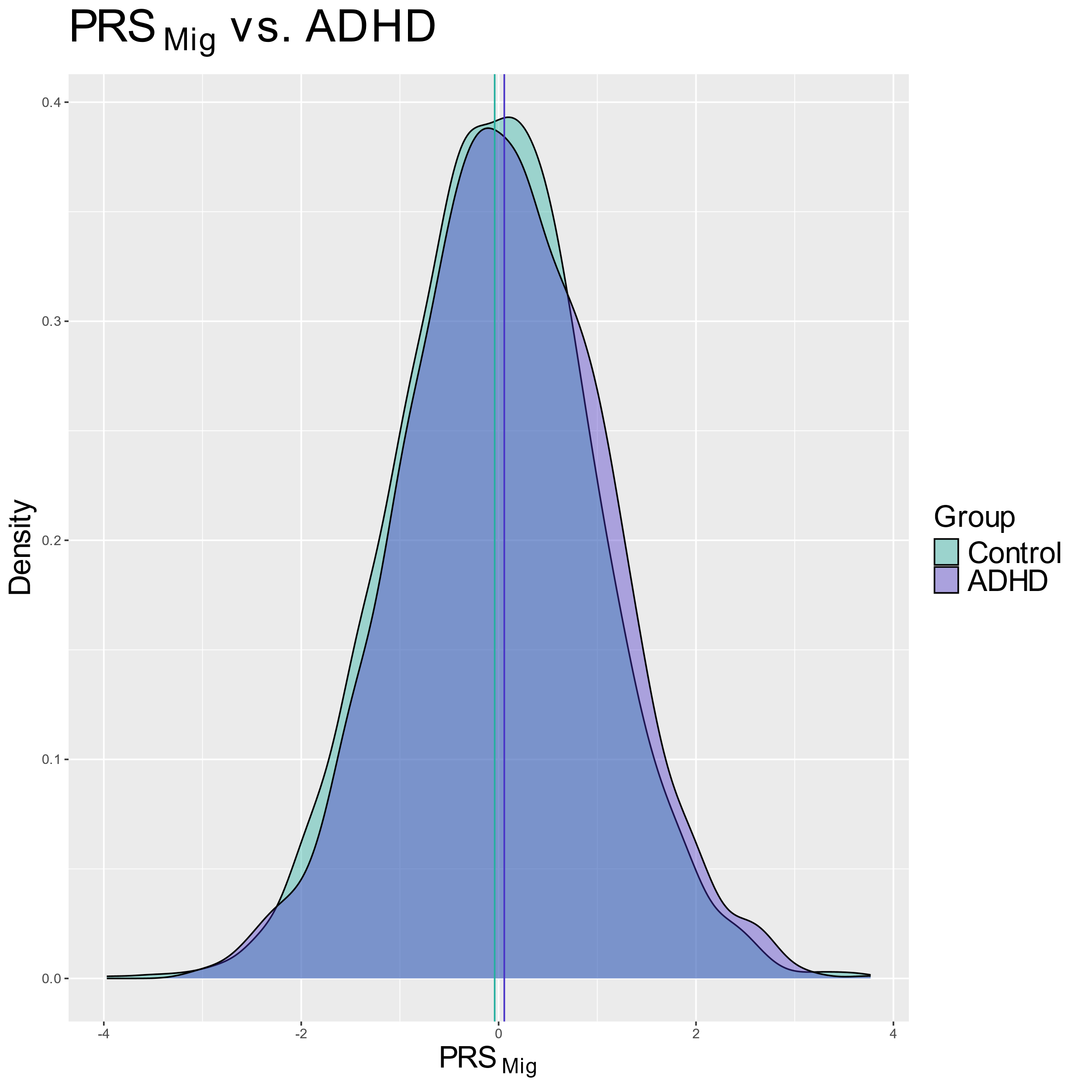

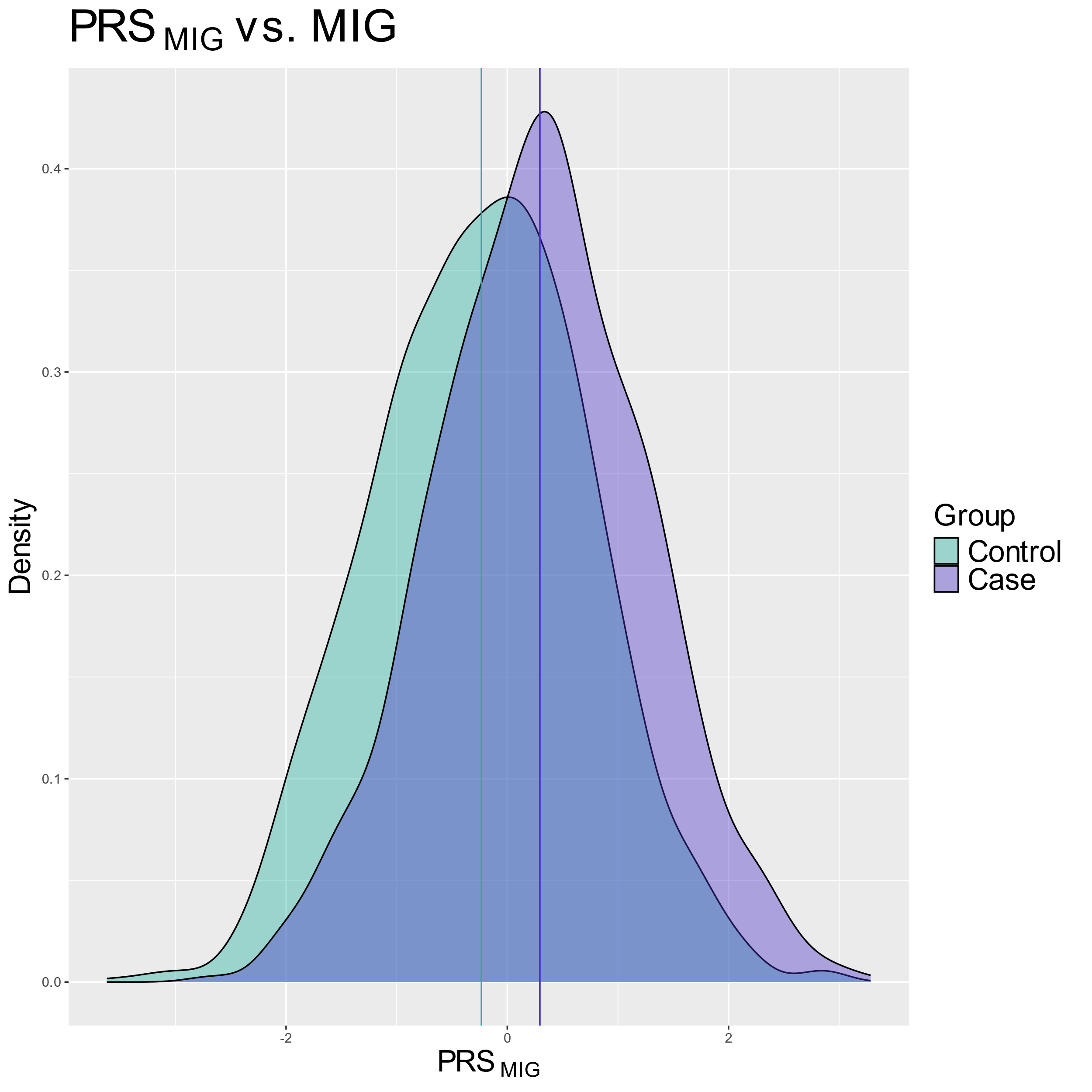

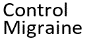

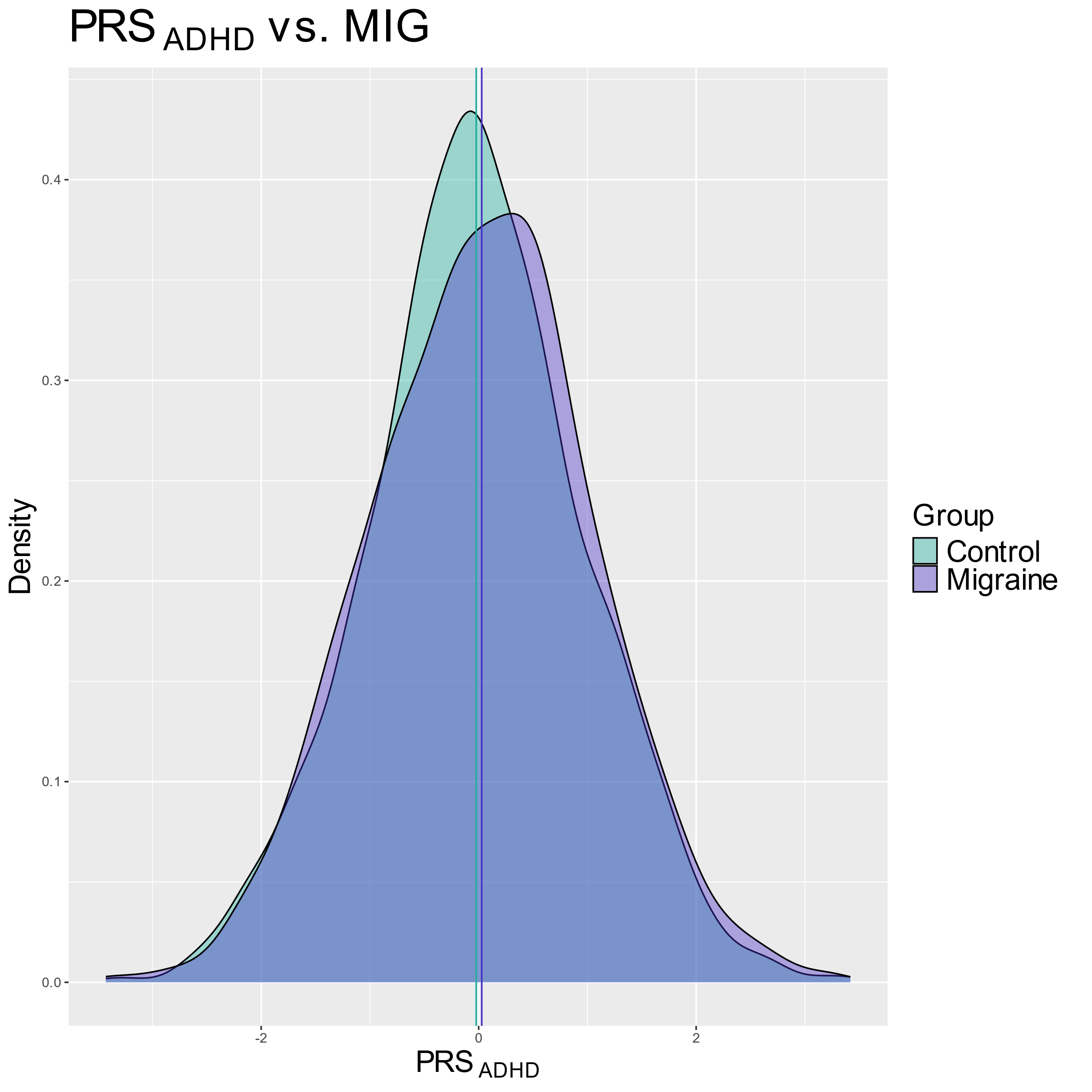

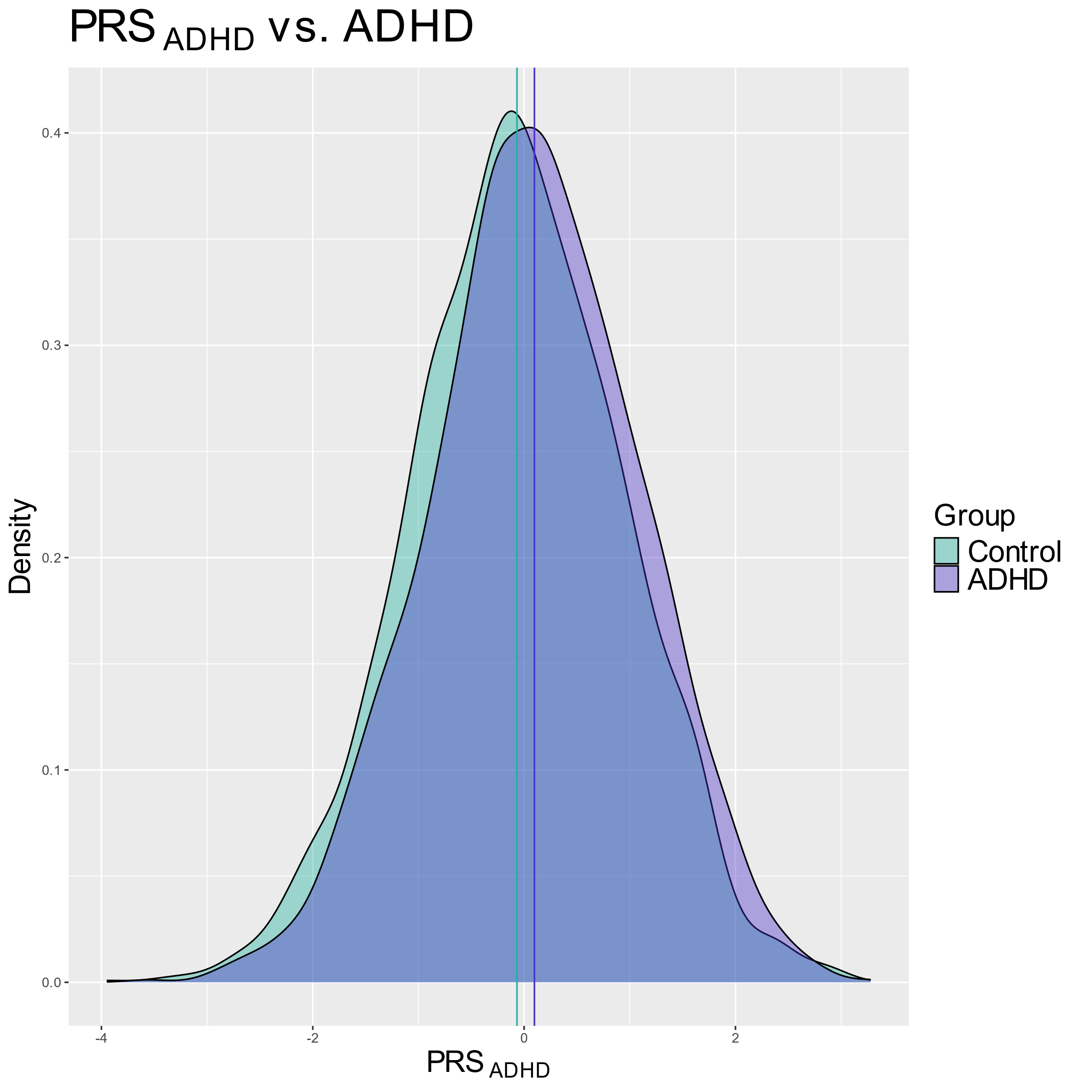

*

*

PRS_MIG_

Supplementary Figure 4. Density plots of ADHD and migraine polygenic risk scores (PRS) in cases and controls samples. Density distributions of PRS for ADHD and migraine are shown separately for cases and controls within each diagnostic sample (ADHD and migraine). **A)** PRS_ADHD_ tested in to case-control ADHD sample. **B)** PRS_MIG_ tested in to case-control ADHD sample**. C)** PRS_ADHD_ tested in to case-control migraine sample **D)** PRS_MIG_ tested in to case-control migraine sample. Asterisks indicate statistically significant differences between groups (P<0.05).

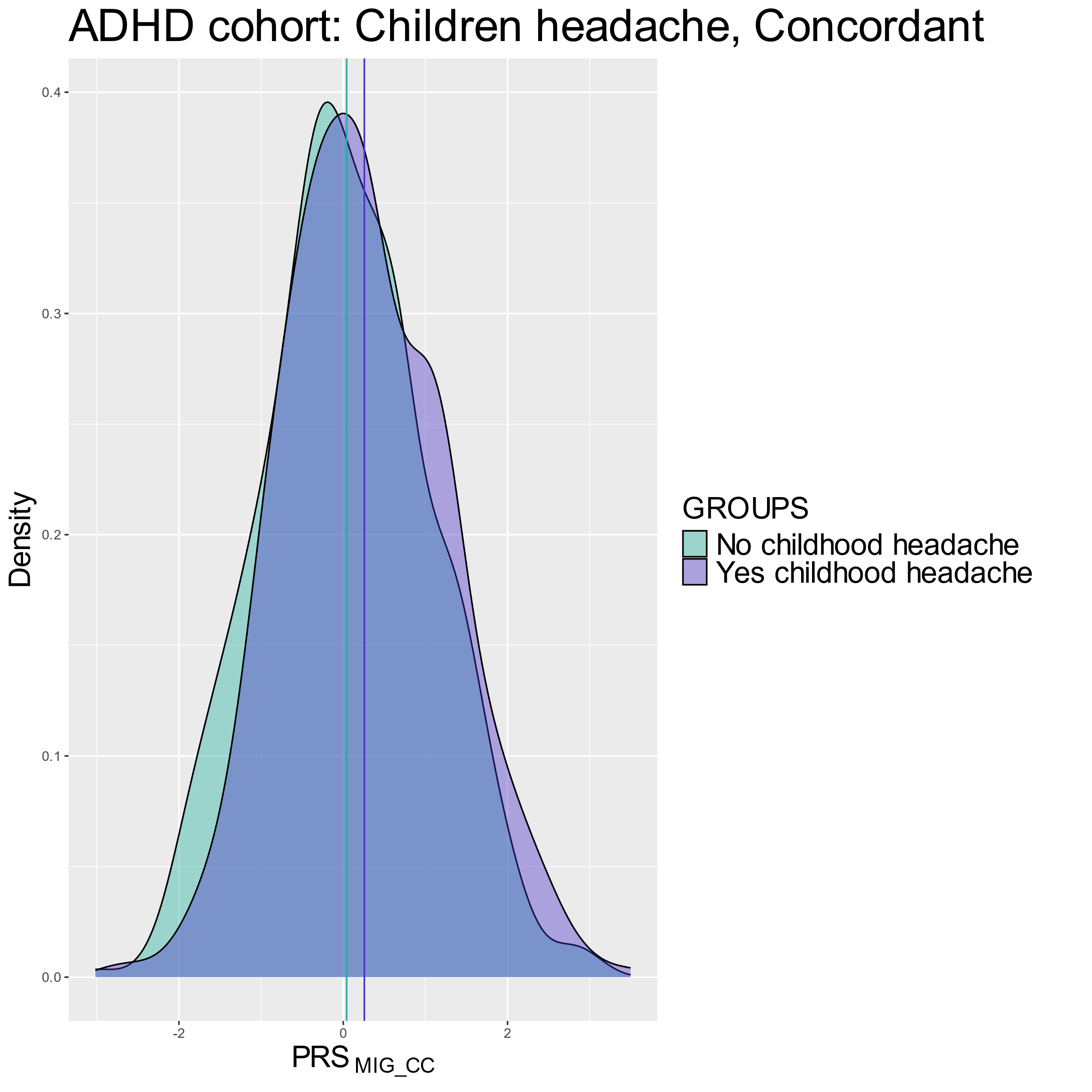

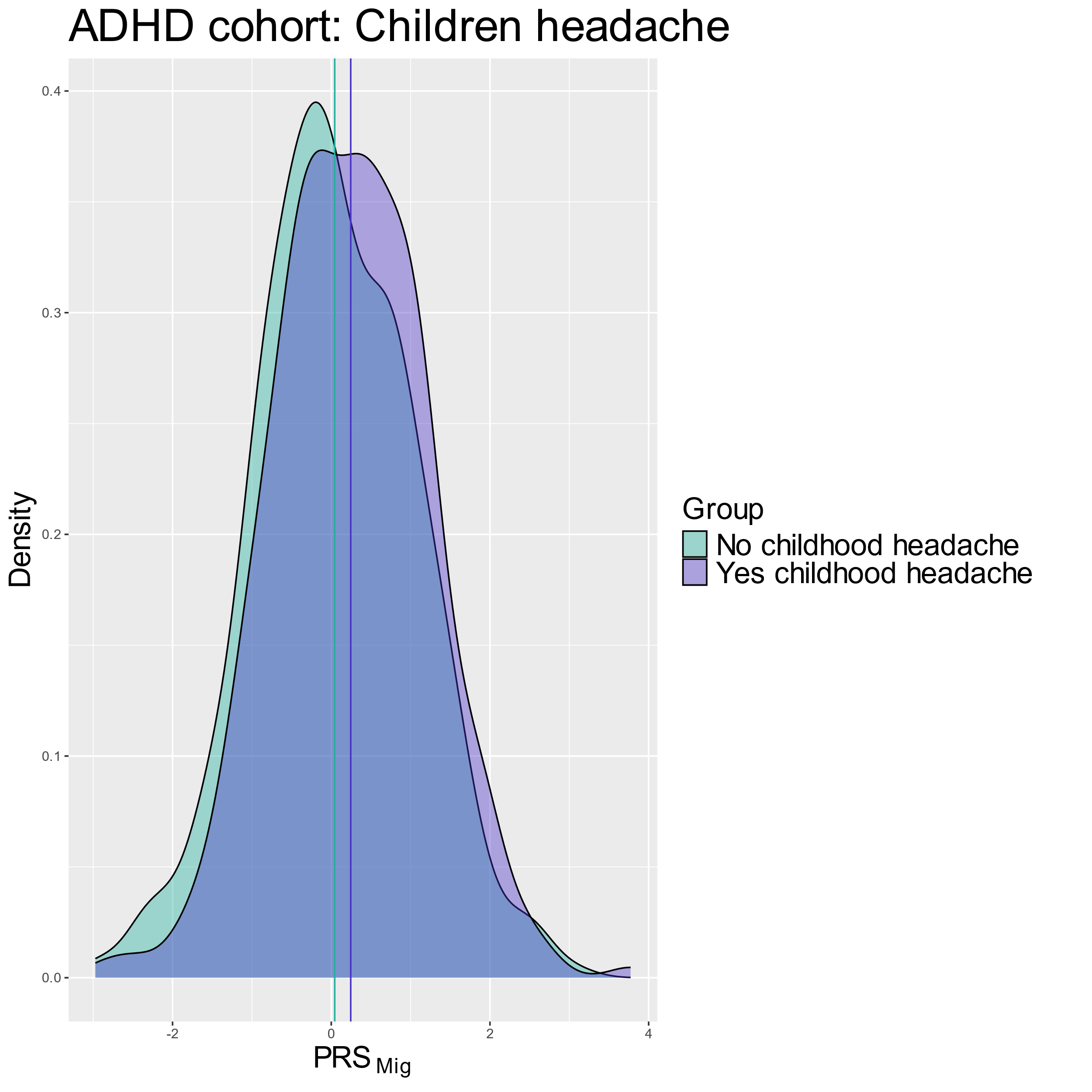

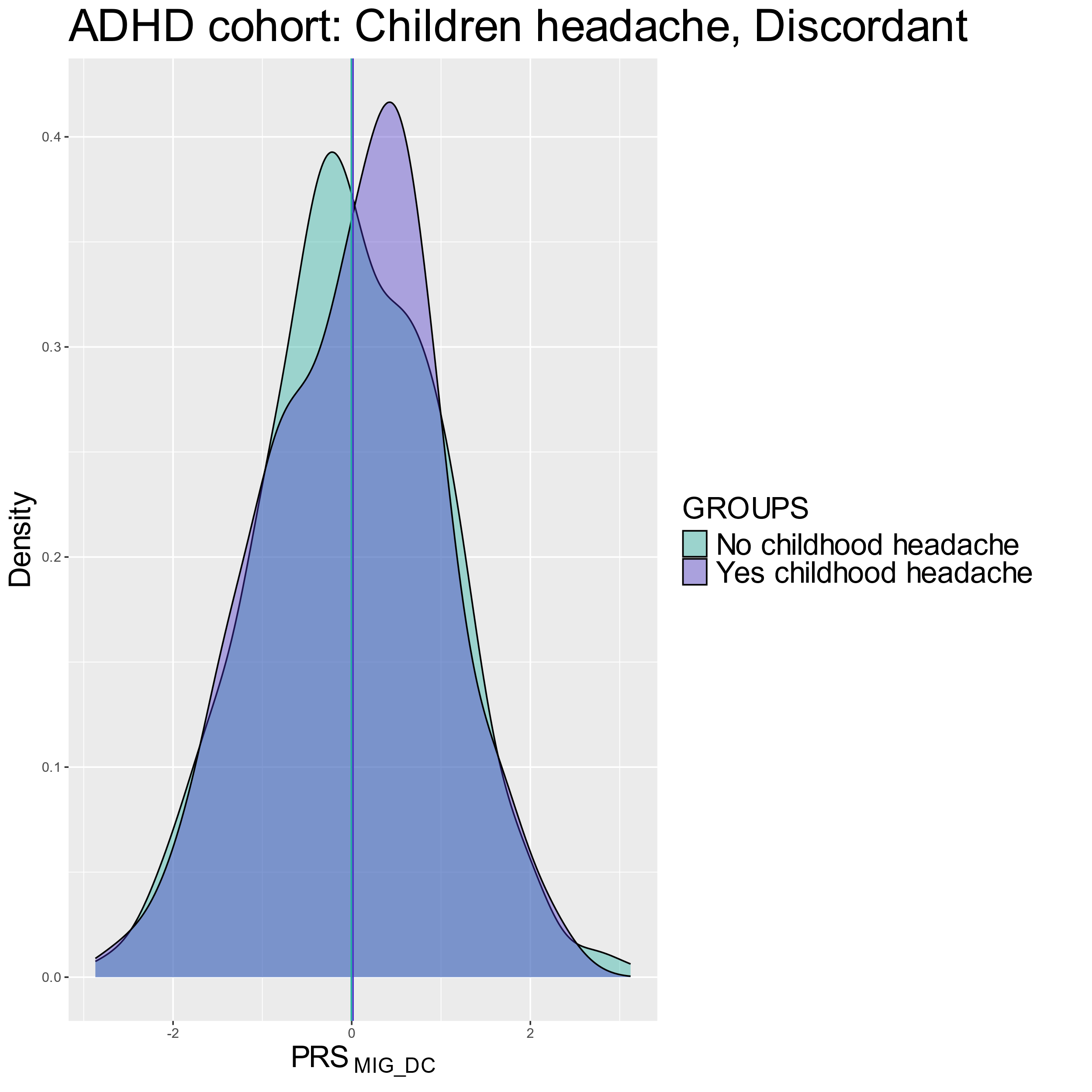

MIG PRS

MIG Discordant PRS

MIG Concordant PRS

*

*

A)

B)

C)

Supplementary Figure 5. Density plots ADHD case-only sample for individuals with and without childhood headache tested for**: A)** migraine PRS **B)** migraine concordant PRS **C)** migraine discordant PRS. Asterisks indicate statistically significant differences between groups (P<0.05).
